# Single-cell analysis of the epigenomic and transcriptional landscape of innate immunity to seasonal and adjuvanted pandemic influenza vaccination in humans

**DOI:** 10.1101/2021.05.24.21253087

**Authors:** Florian Wimmers, Michele Donato, Alex Kuo, Tal Ashuach, Shakti Gupta, Chunfeng Li, Mai Dvorak, Mariko Hinton Foecke, Sarah E. Chang, Sanne E. De Jong, Holden T. Maecker, Robbert van der Most, Peggie Cheung, Mario Cortese, Thomas Hagan, Steven E. Bosinger, Mark Davis, Nadine Rouphael, Shankar Subramaniam, Nir Yosef, Paul J. Utz, Purvesh Khatri, Bali Pulendran

## Abstract

Emerging evidence indicates a fundamental role for the epigenome in immunity. Here, we used a systems biology approach to map the epigenomic and transcriptional landscape of immunity to influenza vaccination in humans at the single-cell level. Vaccination against seasonal influenza resulted in persistently reduced H3K27ac in monocytes and myeloid dendritic cells, which was associated with impaired cytokine responses to TLR stimulation. Single cell ATAC-seq analysis of 120,305 single cells revealed an epigenomically distinct subcluster of monocytes with reduced chromatin accessibility at AP-1-targeted loci after vaccination. Similar effects were also observed in response to vaccination with the AS03-adjuvanted H5N1 pandemic influenza vaccine. However, this vaccine also stimulated persistently increased chromatin accessibility at loci targeted by interferon response factors (IRFs). This was associated with elevated expression of antiviral genes and type 1 IFN production and heightened resistance to infection with the heterologous viruses Zika and Dengue. These results demonstrate that influenza vaccines stimulate persistent epigenomic remodeling of the innate immune system. Notably, AS03-adjuvanted vaccination remodeled the epigenome of myeloid cells to confer heightened resistance against heterologous viruses, revealing its potentially unappreciated role as an epigenetic adjuvant.

## Introduction

Recent research has highlighted a central role for the epigenome in the regulation of fundamental biological processes. The epigenome can maintain particular chromatin states over prolonged periods of time that span generations of cells, thus enabling the durable storage of gene expression information **(Allis and Jenuwein, 2016)**. In the context of the immune system, epigenomic events have been described during hematopoiesis **(Buenrostro et al., 2018; Corces et al., 2016; Farlik et al., 2016)**, generation of immunological memory and exhaustion in T lymphocytes **(Akondy et al., 2017; Satpathy et al., 2019; Youngblood et al., 2017)**, and the development of B and plasma cells **(Barwick et al., 2016; Kulis et al., 2015)**. Recent studies have also revealed that epigenomic changes in monocytes **(Arts et al., 2018; Kleinnijenhuis et al., 2012; Saeed et al., 2014)** and NK cells **(Sun et al., 2011)** imprint a form of immunological memory in the innate immune system **(Netea et al., 2020)**.

The concept of epigenetic imprinting on the innate immune system has acquired a particular significance in the context of vaccination **(Wimmers and Pulendran, 2020)**. Vaccination with live-attenuated BCG has been shown to induce epigenomic changes in monocytes **(Arts et al., 2018; Kleinnijenhuis et al., 2012),** and it has been suggested that such changes result in a durable state of innate activation. However, the extent to which such epigenomic imprinting, observed with BCG vaccination, reflects a more general phenomenon with other vaccines is an open question. Furthermore, the critical parameters that determine vaccination-induced epigenomic imprinting, such as the type of vaccine or adjuvant used, or the impact of the microbiome, are not known. Notably, previous studies identified transcriptional and protein level heterogeneity within monocyte and dendritic cell populations **(Alcántara-Hernández et al., 2017; Arunachalam et al., 2020; Kazer et al., 2020; Schulte-Schrepping et al., 2020; See et al., 2017; Shalek et al., 2014; Villani et al., 2017; Wimmers et al., 2018)**. How this cellular heterogeneity affects epigenomic imprinting during and immune response to a vaccine or to any stimulus is entirely unknown.

Recently, researchers have used systems biology approaches to comprehensively analyze the transcriptional, metabolic, proteomic and cellular landscape in response to vaccination in humans, and identified correlates and mechanisms of vaccine immunity **(Gaucher et al., 2008; Hagan et al., 2015; Kotliarov et al., 2020; Li et al., 2017b; Pulendran et al., 2010; Querec et al., 2009; Team and Consortium, 2017; Tsang et al., 2014; Wimmers and Pulendran, 2020)**. Despite these advances, a comprehensive systems biology assessment of the human epigenomic landscape during an immune response, particularly at the single-cell level, is missing.

In the current study, we used single-cell techniques, including EpiTOF (Epigenetic landscape profiling using cytometry by Time-Of-Flight) **(Cheung et al., 2018)**, single-cell ATAC-seq, and single-cell RNA-seq, to study the epigenomic and transcriptional landscape of immunity to seasonal and pre-pandemic influenza vaccination in humans. We found that vaccination with the trivalent inactivated seasonal influenza vaccine (TIV) induced global changes to the chromatin state in multiple immune cell subsets, which persisted for up to six months after vaccination. These changes were most pronounced in myeloid cells, which demonstrated a transition to inaccessible chromatin in loci targeted by AP-1 transcription factors, and reduced cytokine production in response to TLR stimulation. Single-cell analysis revealed distinct subclusters within the monocyte population that were characterized by differences in AP-1 accessibility. Vaccination with the AS03-adjuvanted H5N1 pre-pandemic influenza vaccine also induced similar epigenomic and functional changes in the innate immune system. Strikingly however, AS03 adjuvanted vaccine also induced a concomitantly enhanced state of antiviral vigilance with increased chromatin accessibility at IRF and STAT loci, and heightened resistance against heterologous viral infection during in-vitro culture.

## Results

### Global epigenomic reprogramming of immune cell subsets after vaccination with TIV

To determine how immunization with TIV affects the epigenomic landscape of the immune system at the single-cell level, we employed EpiTOF to analyze a cohort of 21 healthy individuals aged 18-45 before and after TIV administration (**DataS1**). All subjects received TIV on day 0. Additionally, to determine the impact of the gut microbiota on the epigenomic immune cell landscape, a subgroup of ten subjects received an additional oral antibiotic regimen, consisting of neomycin, vancomycin, and metronidazole, between days -3 and 1 (**Figure 1A**). Our previous work with this cohort had demonstrated that antibiotics administration induced significant changes in the transcriptional and metabolic profiles of peripheral blood mononuclear cells (PBMCs) **(Hagan et al., 2019)**. Therefore, we hypothesized antibiotics administration would induce epigenomic reprogramming of PBMCs. To test this hypothesis, we developed two EpiTOF panels and probed the global levels of 38 distinct histone marks, including acetylation, methylation, phosphorylation, ubiquitination, citrullination, and crotonylation, in 21 distinct immune cell subsets (**DataS2)**. Using EpiTOF, we analyzed PBMCs (**Figure S1A, related to Figure 1**) isolated at day -21 and 0 prior to vaccination, and days 1, 7, 30, and 180 after vaccination. Using a manual gating approach, we detected all major immune cell populations (**DataS2**). While the frequency of immune cell populations did not change significantly between these time points, we observed a trend towards reduced fractions of myeloid cells in some subjects at later time points, and a transient increase in the proportion of pDCs in response to antibiotics treatment (days 0, 1) (**Figure S1B)**, in line with previous observations **(Hagan et al., 2019).** Next, we extracted the histone modification information for each subset and generated a UMAP representation of the epigenomic immune cell landscape (**Figure 1B**). In the UMAP space, lymphoid cells separated from myeloid cells while hematopoietic progenitors (CD34^+^) showed a unique epigenetic pattern distinct from fully differentiated immune cells.

**Figure 1.**
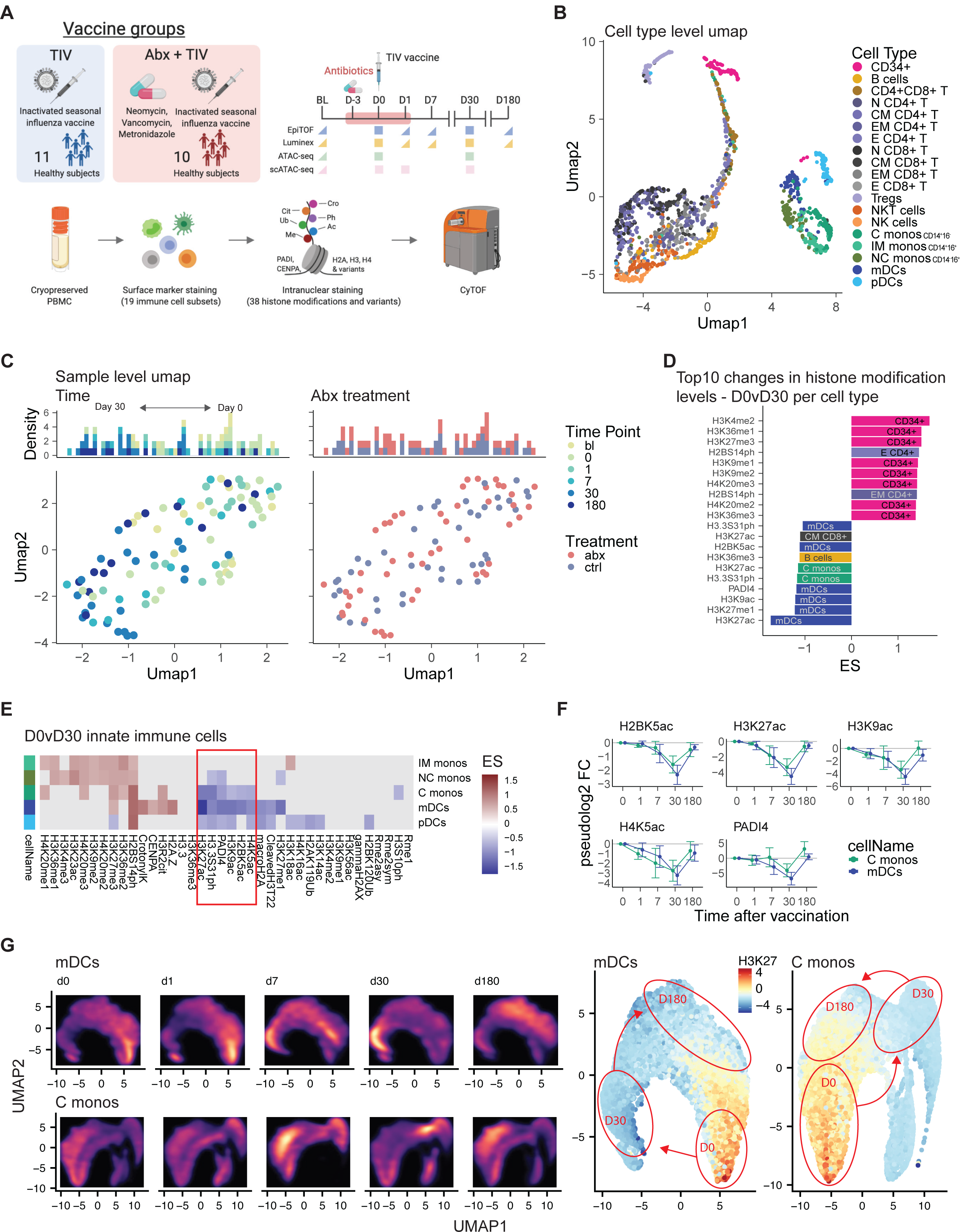
TIV alters the global histone modification profile of immune cells. (A) Study overview. Healthy subjects (n = 21) were vaccinated with TIV at day 0. A subgroup of 11 subjects received an additional oral antibiotic regimen, consisting of neomycin, vancomycin, and metronidazole, between days -3 and 1. The global histone modification profile of PBMCs in these subjects was then determined using EpiTOF. (B) UMAP was used to create a dimensionality-reduced representation of the global histone mark profiles of all immune cell subset. (C) UMAP was used to visualize epigenomic profiles at the sample level. (D, E) The effect size of vaccine-induced changes to the global histone modification profiles at day 30 after vaccination compared to day 0 before vaccination were calculated. D) Top-10 most significantly increased and reduced histone modifications. E) Heatmap showing histone modification changes in innate immune cells. Only changes with an FDR <= 20% are shown. (F) Change in histone modification levels relative to day 0 before vaccination for a set of highly reduced histone modifications in C monos and mDCs. Dots and lines indicate average modification levels, error bars indicate the standard error of mean. (G) Histone modification levels of H2BK5ac, H3K37ac, H3K9ac, H4K5ac, and PADI4 were used to generate a UMAP representation of single monocytes and mDCs at all time points. Left panel: cell density at each time point, right panel: H3K27ac levels in each single cell. Red ellipses indicate the high-density areas at three time points, D0, D30, D180 and correspond to the bright areas in the left panel.

At the sample level, immune cells isolated after vaccination, especially at day 30, were separated in the UMAP space from those collected before vaccination, indicating TIV-induced epigenetic changes (**Figure 1C, left**). Surprisingly, antibiotics status had no measurable impact on histone modification levels and samples from antibiotics-treated and control subjects were intermixed (**Figure 1C, right**). This observation was surprising given the pronounced impact of antibiotics treatment on blood transcriptome and metabolome previously observed in these subjects **(Hagan et al., 2019)**. Rather, we observed changes in the acetylation, methylation, and phosphorylation states of several histone marks in response to vaccination, regardless of exposure to antibiotics (**Figure S1C, D, related to Figure 1**). Thus, we combined both groups for downstream analyses to enhance statistical power. In particular, we detected an increase in several histone methylation marks in CD34^+^ cells and a decrease in multiple acetylation marks in myeloid cells in day 30 samples over baseline (**Figure 1D**). Furthermore, we observed increased H2BS14ph in multiple immune cell subsets at day 30 after vaccination (**Figure S1C**). Elevated H2BS14ph has been shown to occur during apoptosis **(Cheung et al., 2003; Solier and Pommier, 2009; Wen et al., 2010)**. However, we did not observe reduced cell viability at any time point (**Figure S1A, related to Figure 1**), suggesting H2BS14ph functions independent of apoptosis in post-vaccination immune cells. Notably, H2BS14ph is catalyzed by Mst1/STK4, whose immune modulatory role has been reported **(Cho et al., 2019; Li et al., 2017a, 2015; Zhou et al., 2019)**.

### Persistent epigenomic reprogramming in myeloid cells

Classical monocytes and myeloid dendritic cells (mDCs) were characterized by repressed H2BK5ac, H3K9ac, H3K27ac, and H4K5ac at day 30 after vaccination (**Figure 1E**). PADI4, an arginine deiminase catalyzing histone citrullination, was also repressed in these cells. However, global H3R2cit was not reduced in response to PADI4 repression, suggesting potential compensation by other arginine deiminases or localized changes in histone citrullination. Notably, pairwise correlation analysis identified high correlation coefficients between acetylation marks and PADI4 (**Figure S1E, related to Figure 1**), and PADI4 has also been implicated in monocyte development, and inflammation **(Cheung et al., 2018; Liu et al., 2018; Nakashima et al., 1999; Vossenaar et al., 2004)**. Longitudinal analysis demonstrated a time-dependent decrease of the four histone acetylation marks and PADI4, which showed the greatest repression at day 30 and largely returned to baseline levels at day 180 (**Figure 1F**). Blood transcriptomics data obtained from PBMCs of the same subjects at early time points after vaccination **(Hagan et al., 2019)** revealed downregulation of histone acetyltransferases CREBBP/CBP **(Weinert et al., 2018)** and KAT6A **(Voss et al., 2009)** at days 1, 3, and 7 after vaccination over baseline (**Figure S2A, related to Figure 1**). In contrast, various histone deacetylases showed increased expression (**Figure S2A, related to Figure 1**). Moreover, the expression of lysine methyltransferase EZH2 was elevated (**Figure S2A, related to Figure 1**), consistent with increased H3K27me3, an antagonist of H3K27ac, in classical monocytes and myeloid dendritic cells (mDCs) (**Figure 1E**). Epigenomic and transcriptional analysis thus both point towards a, potentially repressive, state of hypoacetylation in myeloid cells after immunization with TIV.

Next, we investigated the TIV-induced epigenomic alterations in myeloid cells at the single-cell level (**Figure 1G**). By performing sub-clustering and UMAP-based dimensionality reduction analysis of mDCs and classical monocytes using the H3K27ac, H2BK5ac, H4K5ac, H3K9ac, and PADI4 marks, we constructed the single-cell histone modification landscape. Importantly, in both cell types, single cells segregated according to vaccination time point with cells at day 0 and 1 clustering together on one side of the 2D space, and cells at day 30 occupying the opposite side (**Figure 1G**). Interestingly, and undetected by the bulk kinetics analysis (**Figure 1F**), cells at day 180 did not return to the baseline position occupied by day 0 cells but assumed an intermediate state (**Figure 1G**), indicating persistent epigenetic alterations that can still be detected up to 6 months after immunization with TIV.

These observations raise the question of how persistent epigenetic changes lasting up to 6 months, can be maintained in monocytes and mDCs, given that these cell types are known to have a relatively rapid turnover of less than 7 days. Recent studies indicate that such persistent changes in circulating myeloid cells are associated with persistent epigenetic changes in the hematopoietic stem and progenitor cell compartment in the bone marrow **(Cirovic et al., 2020; Kaufmann et al., 2018; Mitroulis et al., 2018)**. To determine if this was also evident in the current study, we calculated the epigenomic distance of CD34^+^ cells to a consensus profile of differentiated lymphoid or myeloid cells (**Figure S3A, related to Figure 1**). Interestingly, we detected multiple populations of CD34^+^ cells based on their epigenomic distances with minor populations showing relatively short distances to differentiated immune cells, possibly resembling pre-committed clones (**Figure S3B, related to Figure 1**). After vaccination, the overall distance between CD34^+^ cells and either lymphoid or myeloid cells increased, and the fraction of potentially pre-committed progenitors was greatly reduced (**Figure S3B-D, related to Figure 1**) indicating a potential shift of the stem cell pool towards an immature phenotype after vaccination. At day 180, the distances returned to their pre-vaccination state.

Together, EpiTOF analysis revealed marked epigenetic alterations in immune cells from subjects vaccinated with TIV. In particular classical monocytes and dendritic cells exhibited a concerted reduction in multiple histone acetylation marks and PADI4, that remained detectable 180 days after vaccination.

### TIV induces persistent functional changes in innate immune cells

Given that both histone acetylation and PADI4 activity are associated with gene expression and monocyte function, we wondered whether the observed reduction in these marks at day 30 after TIV had any impact on myeloid cell function. To answer this, we stimulated PBMCs from vaccinated individuals prior to vaccination, or at various time points after vaccination with cocktails of synthetic TLR ligands mimicking bacterial (LPS, Flagellin, Pam-3-Cys) or viral (pI:C, R848) pathogen-associated molecular patterns (**Figure 2A**). After 24h of stimulation, we measured the levels of 62 secreted cytokines in culture supernatants using a multiplexed bead-based assay. To determine whether PBMCs from time points after vaccination showed any alterations in cytokine production, we calculated the relative change in cytokine levels compared to day 0 (**Figure 2B**). Indeed, using hierarchical clustering, we identified a subset of cytokines that displayed a significant reduction at day 30 after vaccination (**Figure 2B** **red box, C**). These cytokines include TNF-a, IL-1b, IL-1RA, IL-12, and IL-10, the monocytic chemokines MCP1, MCP3, ENA78 (CXCL5), and IP-10 (CXCL10), as well as the monocyte growth factor GCSF. Similar to the epigenomic changes, cytokine levels begin to fall around day 1 to 7 after vaccination, reaching a nadir at day 30, and returning to near-baseline levels at day 180 (**Figure 2D**). All of these cytokines were strongly induced by both TLR cocktails (**Figure S4A, related to Figure 2**) and a reduction relative to day 0 was observed in both antibiotics-treated and control subjects **(Figure S4B, related to Figure 2)**.

**Figure 2.**
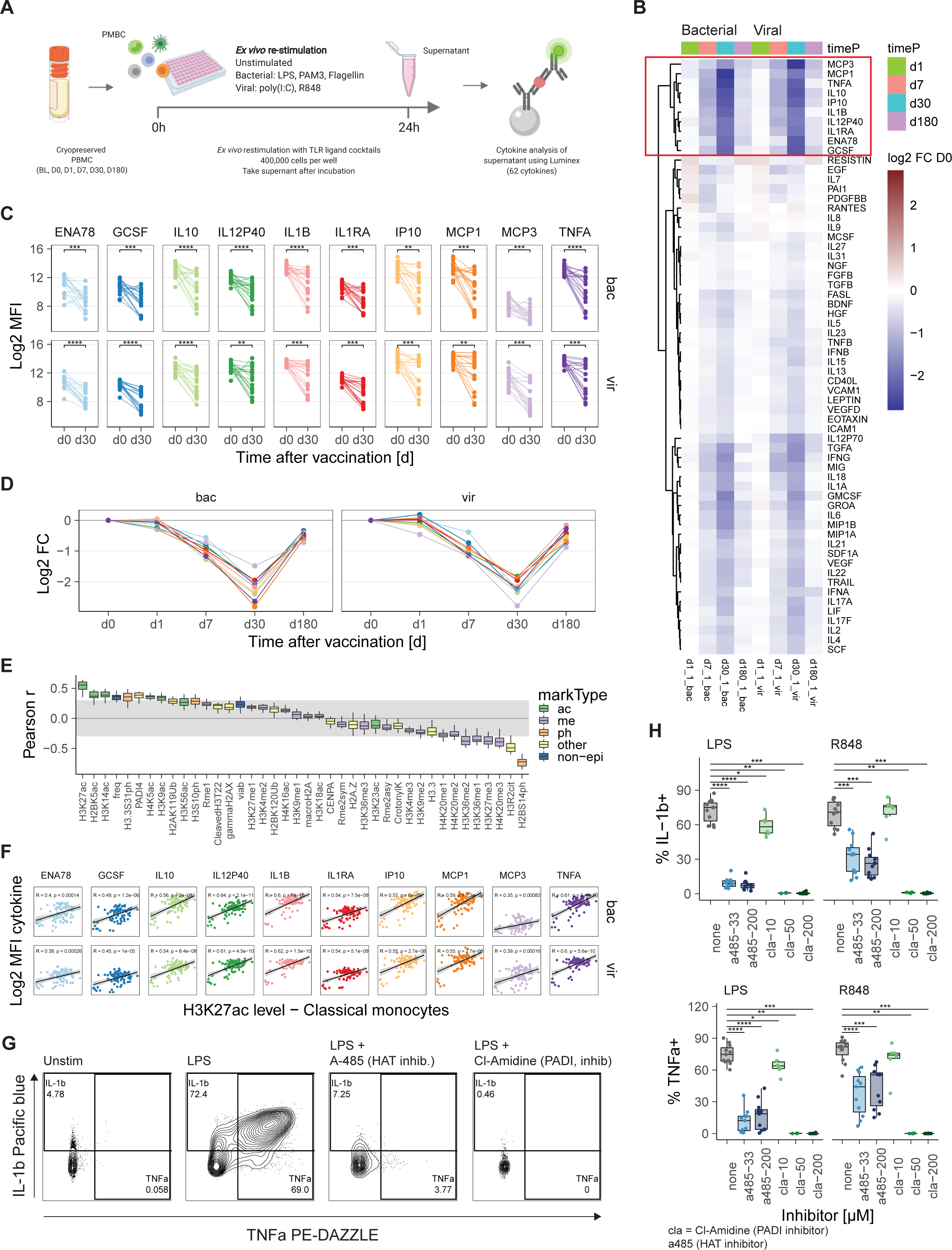
TIV-induced histone modification changes correlate with cytokine production. (A) Schematic overview of experiment. PBMCs from subjects in the EpiTOF experiment were stimulated with three cocktails of synthetic TLR ligands, mimicking bacterial (10 µg/mL Pam3, 25 ng/mL LPS, 300 ng/mL Flagellin) and viral (25 µg/mL pI:C, 4 µg/mL R848) pathogen-associated molecular patterns. After 24h, Luminex was used to measure the cytokine levels in supernatants. (B) Heatmap showing the relative change in cytokine levels at indicated time points compared to day 0. (C) Cytokine levels at day 0 before and day 30 after vaccination for each investigated subject. Wilcoxon signed rank test was used for hypothesis testing. * p <= 0.05, ** p <= 0.01, *** p <= 0.001, **** p <= 0.0001. (D) Change in cytokine levels relative to day 0 for cytokines in C. Dots and lines indicate average. (E, F) Pearson correlation of the cytokine levels of the 10 cytokines in C) with histone modification levels in C monos as well as C mono frequency in PBMCs as determined by EpiTOF and sample viability. (n = 87 samples from all time points) E) Boxplots of correlation coefficients for each cytokine after stimulation with either viral or bacterial cocktail. F) Scatter plots for the indicated histone modifications and cytokines. (G, H,) PBMCs from healthy donors were pre-treated with the pharmacological inhibitors A-485 (P300/CBP), and Cl-Amidine (PADI4) for 2h and subsequently stimulated with either LPS (25 ng/mL) or R848 (4 µg/mL) for 6h. BrefA was added for the last 4h of stimulation. H3K27ac, total H3, IL-1b and TNFa levels were measured using intracellular flow cytometry. G) Gating scheme showing the production of IL-1b and TNFa in C monos after indicated treatment. H) Boxplot summary of the fraction of IL-1b+ or TNFa+ cells in multiple donors. Wilcoxon rank sum test, * p <= 0.05, ** p <= 0.01, *** p <= 0.001, **** p <= 0.0001, n = 4-11

Next, we investigated whether there is a direct relationship between global histone modification levels and TLR-induced cytokine production. We used pairwise correlation analysis to correlate the cytokine levels in a sample with the EpiTOF histone modification levels in classical monocytes and with monocyte frequency in the PBMCs of the same sample and cell viability (**Figure 2E**). Strikingly, the histone acetylation marks previously identified in **Figure 1C**, especially H3K27ac, and PADI4 showed positive correlation with cytokine production (**Figure 2E, F**). In contrast, H2BS14ph and several repressive methylation marks, including H3K27me3 and H4K20me3 **(Cao and Zhang, 2004; Stender et al., 2012)**, were negatively correlated with cytokine production (**Figure 2E**).

Next, we determined if perturbations of global histone acetylation or PADI4 activity affect TLR-induced cytokine secretion. We conducted an ex-vivo stimulation experiment using specific inhibitors for the histone acetyl transferases CBP/p300 (A-485, inhibits acetylation at H3K27, H2BK5, and H4K5**, (Lasko et al., 2017; Weinert et al., 2018)** and PADI4 (Cl-Amidine) followed by stimulation with synthetic TLR ligands. We also used trichostatin A (TSA, enhances histone acetylation via inhibition of HDACs, **(Kim and Bae; Yoshida et al., 1990)** as a positive control for detecting histone acetylation. Using flow cytometry, we assessed expression of H3K27ac and the intracellular accumulation of IL-1b and TNFa. As expected, treatment with the histone acetyl transferase inhibitor A-485 led to a concentration-dependent decrease in global histone H3K27ac levels in classical monocytes while treatment with the HDAC inhibitor TSA generated a concentration-dependent increase (**Data not shown**). Furthermore, treatment with the PADI4 inhibitor Cl-Amidine led to similar reductions in H3K27ac (**Data not shown**) in line with the strong correlation of PADI4 and H3K27ac levels in EpiTOF (**Figure S1E**) and the previously observed ability of PADI4 to regulate CBP/P300 **(Lee et al., 2005)**. Notably, none of these inhibitors had an effect on cell viability (**Data not shown**). Next, we asked whether inhibition of CBP/P300 and PADI4 has an impact on cytokine production. Indeed, treatment with A-485 led to a major diminution in the frequency of IL-1b and TNFa positive monocytes after stimulation with LPS or R848 (**Figure 2G, H**). Cl-Amidine treatment, strikingly, led to a complete abrogation of cytokine production in these cells (**Figure 2H**). Together, these results demonstrate reduced innate immune cell functionality after TIV vaccination that is correlated with hypoacetylation and reduced PADI4 in monocytes. Furthermore, altered HAT and PADI4 catalytic activities directly impact cytokine secretion by monocytes.

### Vaccination against seasonal influenza induces reduced chromatin accessibility of AP-1 targeted loci in myeloid cells

To gain greater insight into the epigenomic changes induced by vaccination, we conducted ATAC-seq analysis of FACS purified innate immune cell subsets before and after vaccination (**Figure 3A**). After preprocessing, we retained a high-quality dataset of 51 unique samples (**DataS3).** To identify the molecular targets of the TIV-induced epigenomic changes, we determined genomic regions with significantly changed chromatin accessibility at day 30 after vaccination compared to day 0. Overall, we detected more than 10,000 differentially accessible regions (DARs) in CD14^+^ monocytes and ∼ 4,500 DARs in mDCs, while pDCs showed only minor changes (**Figure 3B**). In line with reduced histone acetylation levels detected by EpiTOF, the majority of DARs in monocytes and mDCs showed a reduction in chromatin accessibility indicating reduced gene activity (**Figure 3B**). In contrast, comparing samples from day - 21 before antibiotics treatment and day 0 during antibiotics treatment showed no profound change in chromatin accessibility (**Figure S5A, related to Figure 3**) and D0vD30 DARs correlated well between antibiotics-treated and control subjects (**Figure S5B, related to Figure 3**). Among the top 200 DARs in CD14^+^ monocytes, we identified many immune-related genes with reduced accessibility, including several cytokines and chemokines and their associated receptors (IL18, CCL20, CXCL8, CXCL3, IL4R, IL6R-AS1), pathogen recognition receptors (CLEC5A, CLEC17A), and adhesion molecules (CD44, CD38) (**Figure 3C**). We also observed reduced accessibility in regions coding for molecules associated with Ras-MAPK-AP-1 signaling (RAP2B, ETS1, MAP3K8, DUSP5) (**Figure 3C**). Importantly, genomic loci associated with seven of the ten cytokines with reduced post-vaccination levels during *ex-vivo* stimulation showed reduced accessibility (**Figure 2**, **Figure 3C, right panel**). Interestingly, these reduced DARs were predominantly located in non-promoter regions (**Figure 3C**) suggesting the involvement of distal regulatory elements such as enhancers. Pathway analysis followed by network analysis of all DARs in CD14^+^ monocytes revealed two major biological themes: TLR and cytokine signaling, and genome rearrangement (**Figure 3D**). The TLR and cytokine cluster was dominated by pathways with mostly reduced chromatin accessibility while terms in the genome rearrangement cluster were mixed. Notably, DARs associated with signaling pathways around Ras and MAPK signaling were enriched as well.

**Figure 3.**
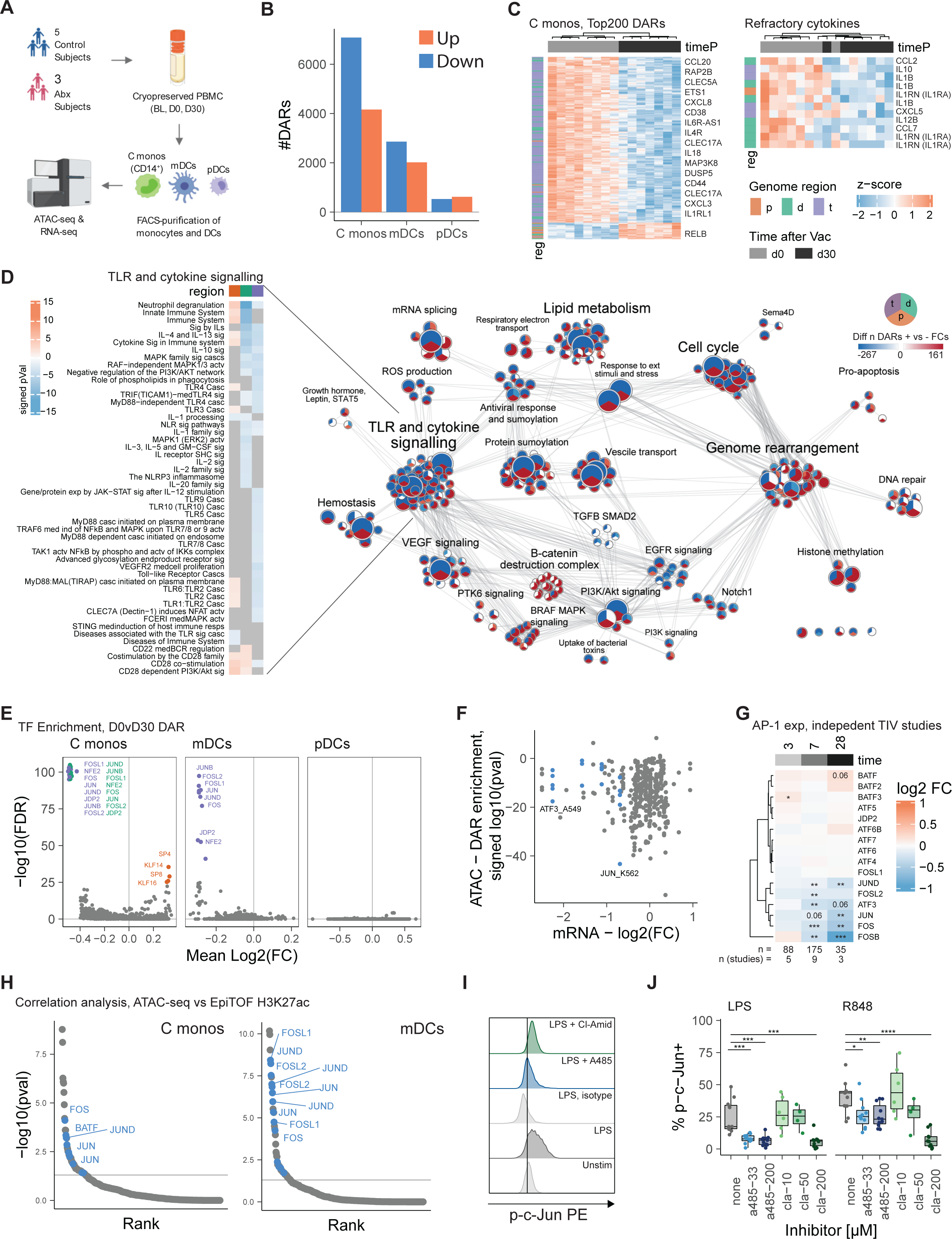
TIV induces reduced chromatin accessibility in immune response genes and AP-1 controlled regions. (A) Schematic overview of the experiment. C monos, mDCs, and pDCs were isolated from PBMCs of vaccinated subjects (n = 8) at day -21 (BL), 0 and 30 using FACS. ATAC-seq and RNA-seq were used to analyze the chromatin accessibility landscape as well as the transcriptional landscape in these cells. (B) Differentially accessible chromatin regions (DARs) at day 30 after vaccination compared to day 0 before vaccination were identified using DESeq2. Pval <= 0.05. (C) Heatmap representation of the normalized accessibility at the top 200 as well as cytokine-associated DARs in C monos for each analyzed sample. p: promoter -2000 bp to +500 bp; d: distal -10kbp to +10kbp – promoter; t: trans < -10kbp or > +10kbp. (D) Network representation of gene set enrichment analysis of DARs in C monos using the Reactome database. Only significantly enriched terms (p <= 0.05) are shown. Color indicates whether majority of enriched regions showed enhanced (red) or reduced (blue) accessibility. Heatmaps show signed –log10(pval) for significantly enriched terms in highlighted clusters. (E) Motif-based overrepresentation analysis of transcription factor binding sites in DARs at day 30 compared to day 0. (F) Scatter plot showing the change in TF gene expression (x-axis) plotted against the enrichment in DARs for selected transcription factors in the Encode database. Blue color indicates AP-1 members with significantly reduced expression. (G) The change in gene expression of AP-1 family members was calculated using bulk transcriptomics data from 3-9 independent flu vaccine trials previously conducted. Heatmap indicates average log2 fold change in gene expression over all trials. N indicates subject and study number at each time point. Wilcoxon signed rank test, * p <= 0.05, ** p <= 0.01, *** p <= 0.001. (H) DARs in indicated cell type were correlated with H3K27ac levels as measured by EpiTOF and DARs with correlation coefficient > .5 were analyzed for transcription factor target gene enrichment using the Encode database. Blue color indicates significantly changed AP-1 members. (I, J) PBMCs from healthy donors were pre-treated with the pharmacological inhibitors A-485 (P300/CBP), and Cl-Amidine (PADI4) for 2h and subsequently stimulated with either LPS (25 ng/mL) or R848 (4 µg/mL) for 6h. BrefA was added for the last 4h of stimulation. Phospho-c-Jun levels were measured using intracellular flow cytometry. Histogram showing the level of phospho-c-Jun in C monos in the indicated conditions (I). Box plot summary of the fraction of phospho-c-Jun positive cells in C monos (J). Wilcoxon rank sum test, * p <= 0.05, ** p <= 0.01, *** p <= 0.001, **** p <= 0.0001, n = 4-11

Next, to identify regulatory patterns, we determined whether the identified DARs in each cell type were enriched for transcription factor (TFs) binding motifs. Indeed, we observed an enrichment for bZIP TFs of the AP-1 family including c-Jun and c-Fos in DARs of monocytes and mDCs, and on average DARs carrying such a motif showed a reduction in chromatin accessibility after vaccination, especially in non-promoter regions (**Figure 3E**). Gene set analysis of the DARs in classical monocytes further confirmed this finding and showed strong enrichment for target genes of c-Jun in DARs with reduced accessibility at day 30 (**Figure 3F**). In addition, we observed reduced expression of several AP-1 family members, including c-Jun, at day 30 after vaccination (**Figure 3F**). Using bulk transcriptomics from previous systems vaccinology studies **(Barrett et al., 2013; Mohanty et al., 2015; Nakaya et al., 2011, 2015; Thakar et al., 2015; Tsang et al., 2014)**, we confirmed the reduced c-Jun expression in up to 9 independent flu vaccine studies and additionally identified a reduction in expression of the AP-1 members JUND, ATF3, FOS, FOSL2, and FOSB (**Figure 3G**). Similar to the histone acetylation changes, the reduction in AP-1 TF expression was first detected at day 7 after vaccination and was most pronounced at day 28 (**Figure 3G**).

Based on this observation, we asked whether the observed reduction in AP-1 accessibility is related to the reduced levels of histone acetylation. To test this, we correlated the normalized accessibility levels of all genomic regions in every sample to histone mark levels in classical monocytes or mDCs isolated from the same sample. Using enrichment analysis on the highly correlated peaks (cor coef > 0.5), we identified a significant enrichment of target genes for multiple AP-1 family members, including c-Fos and c-Jun (**Figure 3H**).

These findings suggest the possibility of a causal link between reduced histone acetylation/PADI4 and reduced AP-1 accessibility. Indeed, previous studies described a direct physical interaction and functional co-dependence between AP-1 and the histone acetyl transferases CBP/P300 **(Arias et al., 1994; Kamei et al., 1996; Zanger et al., 2001)**. To investigate whether AP-1 activity and histone acetylation are also functionally linked in classical monocytes, we conducted an ex-vivo stimulation experiment using the same specific inhibitors of histone acetylation and PADI activity as in Figure 2. To gauge AP-1 activity, we used a monoclonal antibody specific for the activated form of c-Jun, phosphorylated at serine 73. While treatment with LPS or R848 alone induced a robust upregulation of p-c-Jun that can be readily detected by flow cytometry (**Figure 3I, J)**, pre-treatment with A-485 or Cl-Amidine, which lead to reduced histone acetylation, abolished c-Jun activation completely (**Figure 3I, J**).

Together, these results demonstrate that reduced chromatin accessibility in classical monocytes and mDCs after vaccination with TIV. Reduced accessibility is primarily found in regions that are associated with TLR- and cytokine-related genes and regions that carry the AP-1 TF binding motif. Furthermore, HAT/PADI activity is linked to AP-1 activation.

### Single-cell epigenomic and transcriptional landscape of the innate response to seasonal influenza vaccination

Previous studies using transcriptomics and proteomics approaches detected ample heterogeneity within monocyte and dendritic cell populations at steady state **(Alcántara-Hernández et al., 2017; Guilliams et al., 2018; See et al., 2017; Villani et al., 2017)**. However, it is unclear how this heterogeneity affects the epigenomic landscape in these cells and their response to vaccination. To answer this question, we used scATAC-seq and scRNA-seq and constructed the single-cell landscape of the innate immune response to TIV at the epigenomic and transcriptional level. PBMCs from vaccinated individuals were isolated at day 0, 1, and 30, then enriched for DC subsets using flow cytometry and analyzed using droplet-based single-cell gene expression and chromatin accessibility profiling (**Figure 4A**). After initial pre-processing, we obtained chromatin accessibility data from 62,101 cells with an average of 4,126 uniquely accessible fragments. These cells displayed the canonical fragment size distribution and showed high signal-to-noise ratio at transcription start sites (**data not shown**). Using UMAP representation and chromVAR TF deviation patterns, we generated an epigenomic map of the innate immune system and identified clusters for all major innate immune cell subsets, including classical and non-classical monocytes, mDCs, and pDCs (**Figure 4B, DataS3**). In parallel, we used the scRNA-seq data to construct a gene expression map. After pre-processing, we retained 34,368 high-quality transcriptomes with an average of 2,477 genes and 8,951 unique transcripts detected per cell. UMAP representation in combination with clustering allowed us to identify all major innate immune cell subsets (**DataS3**). These subsets were found at all vaccine time points and in sample of all subsets (**DataS3**).

**Figure 4.**
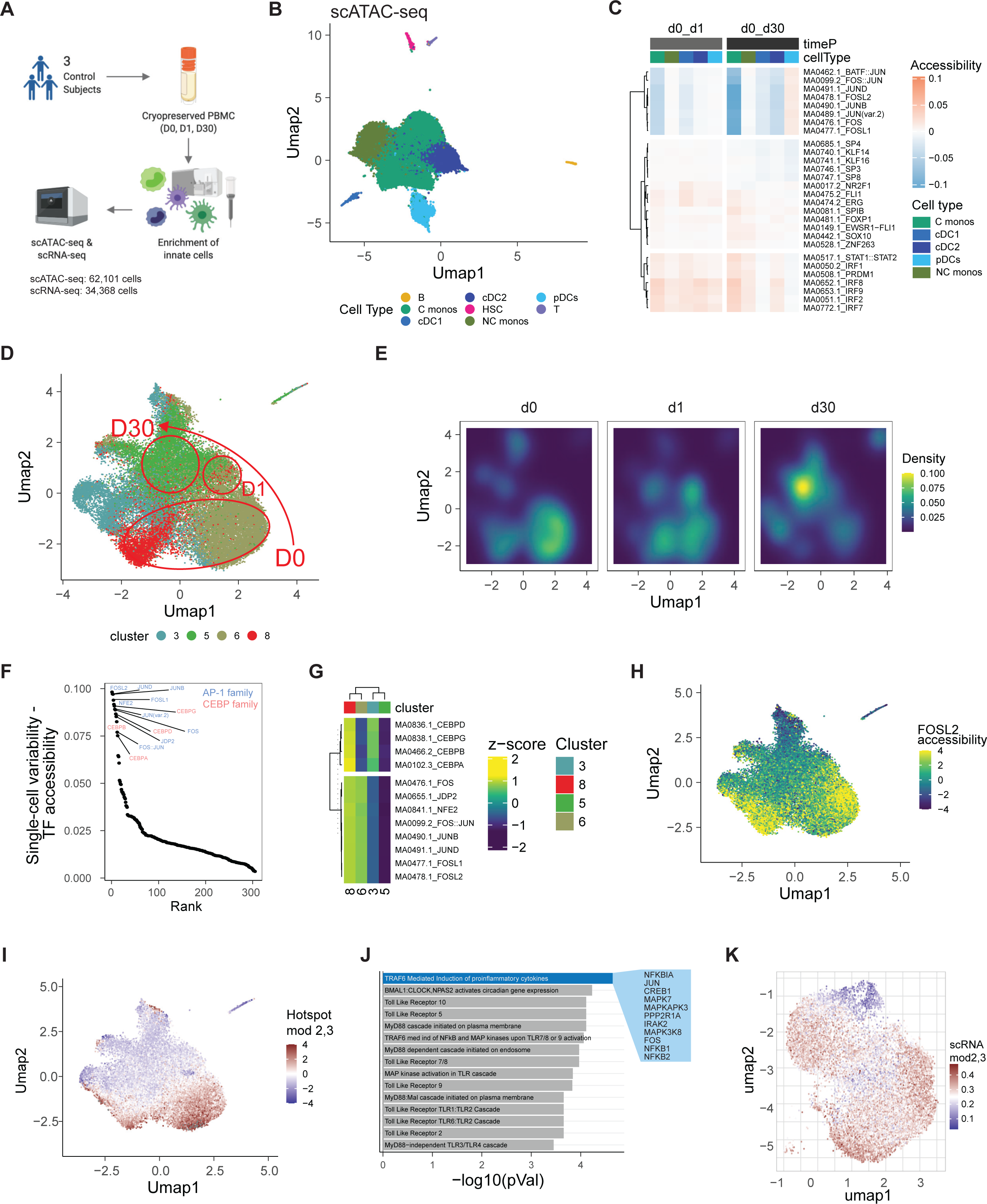
Heterogeneity within monocyte population drives TIV induced epigenomic changes. (A) Schematic overview of the experiment. Innate immune cells were isolated from PBMCs of 3 vaccinated subjects at days 0, 1, and 30, and analyzed using scATAC-seq and scRNA-seq. (B) UMAP representation of scATAC-seq landscape after pre-processing and QC filtering. (C) Heatmap showing the difference in chromatin accessibility at the indicated time points for the top5 transcription factors per subset. (D) UMAP representation of epigenomic subclusters within the classical monocyte population. (E) Density plot showing the relative contribution of different epigenomic subclusters to the total monocyte population at a given vaccine time point. (F) Variability in TF accessibility within the monocyte population. Value indicates range of accessibility values in all single monocytes. (G) Heatmap showing the difference in chromatin accessibility between monocyte subclusters subset. (H) UMAP representation of monocyte subclusters showing differences in AP-1 accessibility. (I) UMAP representation of monocyte subclusters showing difference in accessibility at Hotspot module 2,3 gene loci. (J) Enrichment analysis of genes associated with loci in Hotspot module 2,3. (K) UMAP representation of the transcriptional landscape of single monocytes. Color indicates expression of genes associated with Hotspot modules 2,3.

First, we used chromVAR to determine the TIV-induced changes in TF chromatin accessibility. AP-1 accessibility was strongly reduced at day 30 after vaccination in classical monocytes and mDCs (both cDC1 and cDC2) (**Figure 4C**) confirming our findings with bulk ATAC-seq (**Figure 3**). In addition, using the single-cell dataset, we observed that the reduction in AP-1 accessibility starts early, at day 1 after vaccination (**Figure 4C**) suggesting that the TIV-induced epigenomic reprogramming is imprinted during the acute phase of the vaccine response. At the gene level, we observed a reduction in the expression of multiple AP-1 members including ATF3, JUND, JUNB, FOS, and FOSL2 (**Data not shown**).

Next, we determined the impact of cellular heterogeneity on TIV-induced epigenomic changes. Sub-clustering analysis of classical monocytes revealed the presence of four distinct populations based on chromatin accessibility (**Figure 4D, DataS3**) with different temporal patterns (**Figure 4E**): while clusters 6 and 8 dominated the classical monocyte pool at day 0, most cells at day 30 belonged to cluster 5 **(Figure 4D, E)**. Notably, the observed heterogeneity between the classical monocyte populations was driven by differences in AP-1 and, to a lesser extent, CEBP accessibility (**Figure 4F**). While the dominating clusters at day 0 (cluster 6 and 8) were high in AP-1 accessibility, cells in cluster 5, which was predominantly found at day 30, were low in AP-1 (**Figure 4G, H**). Cells in cluster 3 exhibited intermediate AP-1 and CEBP accessibility (**Figure 4G**) and their relative abundance was stable throughout vaccination (**Figure 4E**). Using Hotspot **(DeTomaso and Yosef, 2020),** we determined a set of genomic regions underlying the observed heterogeneity (**Figure 4I**). This set was enriched for regions associated with the production of proinflammatory cytokines and TLR signaling (**Figure 4J**), and included regions associated with the AP-1 members FOS and JUN, multiple MAP kinases, and NFKB. Importantly, cells with high AP-1 accessibility using the motif-based chromVAR analysis also displayed high accessibility at these regions coding for inflammatory genes (**Figure 4H, I)**. Finally, using the scRNA-seq dataset, we determined the cellular heterogeneity at the transcriptional level. Although genes in the Hotspot module varied in their expression between single classical monocytes, this heterogeneity was less distinct compared to the epigenomic landscape (**Figure 4K**).

Together, these findings demonstrate that AP-1 accessibility drives epigenomic heterogeneity within the pool of classical monocytes and defines epigenomic subclusters. Importantly, changes in the relative abundance of these epigenomic subclusters underly the observed global reduction in AP-1 accessibility after vaccination and a population of AP-1 low, seemingly less inflammatory cells dominated the monocyte pool at day 30.

### AS03 adjuvanted H5N1 influenza vaccine induces reduced chromatin accessibility of AP-1 loci in myeloid cells

The effects of the inactivated seasonal influenza vaccination in inducing reduced chromatic accessibility of AP-1 loci, and reduced H3K27ac and refractoriness to TLR stimulation by myeloid cells, was unexpected and seemingly at odds with prior work on the live attenuated BCG vaccine showing enhanced and persistent innate responses to vaccination, termed “trained immunity” **(Arts et al., 2018)**. This raised the possibility that the live BCG vaccine delivered potent adjuvant signals that stimulated persistent epigenomic changes in myeloid cells, whereas the seasonal influenza vaccine, devoid of an adjuvant, was unable to stimulate trained immunity and instead induced a form of trained tolerance. We hypothesized that the addition of an adjuvant to an inactivated influenza vaccine would induce enhanced and persistent innate responses. We used AS03, a squalene-based adjuvant containing alpha-tocopherol that induces strong innate and adaptive immune responses and is included in the licensed H5N1 avian influenza vaccine (**Garçon et al., 2012; Khurana et al., 2018**). We investigated the effect of AS03 on the epigenomic immune cell landscape in a cohort of healthy individuals that were vaccinated with an inactivated split-virion vaccine against H5N1 influenza administered with or without AS03 (**Figure 5A**). The vaccine was administered in a prime-boost regimen and individuals received injections at day 0 and day 21.

**Figure 5.**
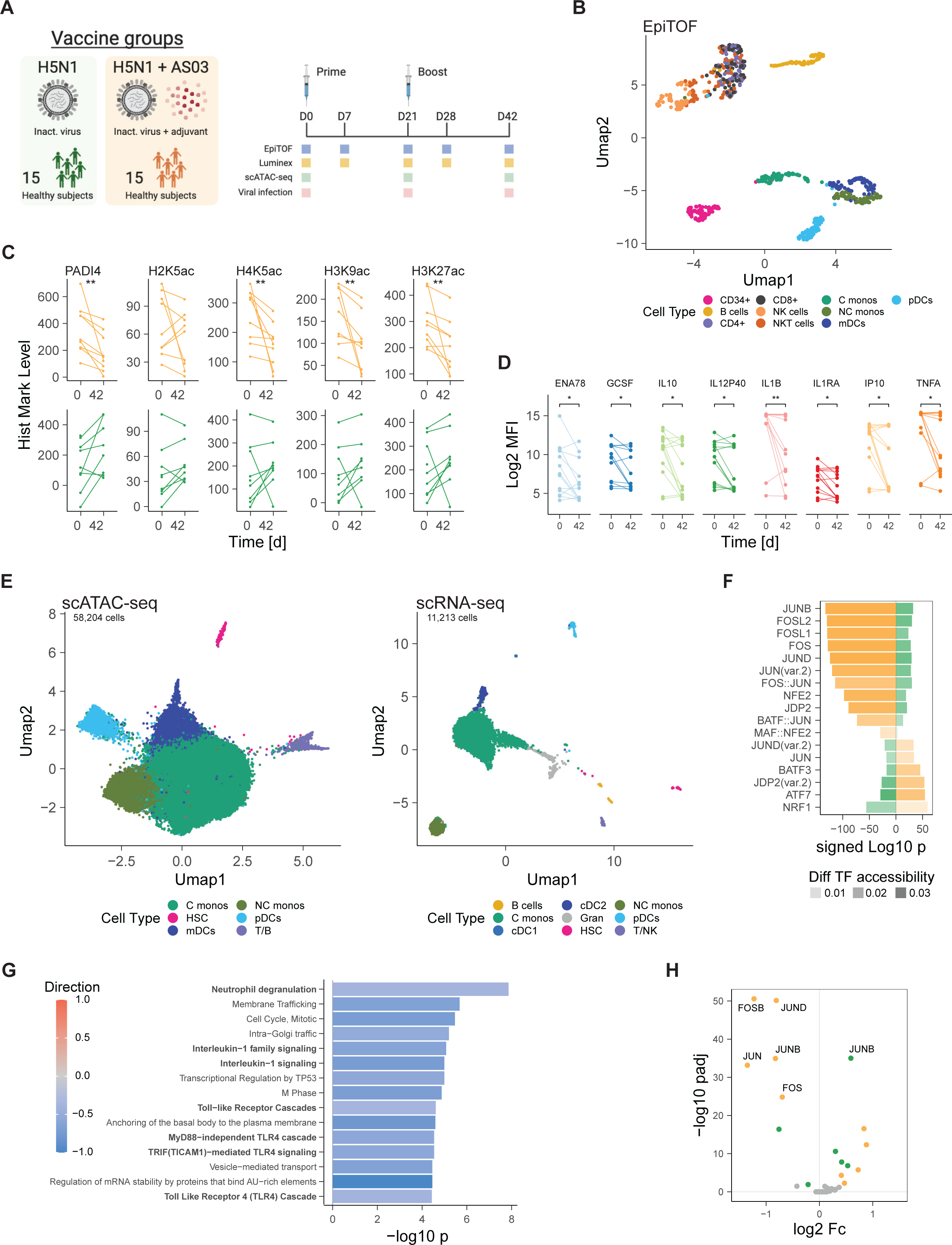
H5N1+AS03 induces repressive epigenomic state akin to TIV. (A) Schematic overview of experiment. Healthy subjects were vaccinated with H5N1 or H5N1 with Adjuvant System 03 (H5N1+AS03) at day 0 and 21. Innate immune cells were isolated from PBMCs at day 0, 21, and 42 and analyzed using scATAC-seq and scRNA-seq (n = 2/2). Ex-vivo TLR stimulation and EpiTOF analysis were conducted on PBMCs at days 0, 7, 21, 28, 42. (n = 9-13/9-13) (B) UMAP representation of EpiTOF landscape. (C) Histone modification levels in classical monocytes at day 0 and day 42 as measured by EpiTOF. (D) Cytokine levels in supernatant of TLR-stimulated PBMCs at day 0 and day 42 after vaccination with H5N1+AS03. (C, D) Wilcox signed rank test; *p <= 0.05, ** p <= 0.01; EpiTOF: n = 9/9, Luminex: n = 13/13. (E) UMAP representation of scATAC-seq (left) and scRNA-seq (right) landscape after pre-processing and QC filtering. (F) Change in accessibility of detected AP-1 family TFs in classical monocytes. Color indicates whether cells are derived from subjects vaccinated with H5N1 (green) or H5N1+AS03 (orange). (G) Overrepresentation analysis of significantly different DARs in classical monocytes using the Reactome database. Color indicates whether enriched genes were predominantly up- or down-regulated. (H) Volcano plot showing changes in expression of AP-1 TF genes in classical monocytes at D42 compared to D0.

First, we asked how the presence of AS03 would affect the vaccine-induced chromatin mark changes observed after vaccination with TIV. Using EpiTOF, we analyzed PBMC samples from 18 vaccinated subjects (9 H5N1, 9 H5N1+AS03) at day 0, 7, 21, 28, and 42 and constructed the histone modification profile landscape (**Figure 5B, Figure S6A, related to Figure 5**). Comparing histone modification profiles at day 0 with day 42, we, unexpectedly, observed that vaccination with H5N1+AS03 induced a significant reduction in H3K27ac, H4K5ac, H3K9ac, and PADI4 in classical monocytes, four of the five highly correlated marks associated with myeloid reprogramming after TIV (**Figure 5C**). In contrast, vaccination with H5N1 alone did not induce significant changes in these chromatin marks. In line with these findings, we also observed in the H5N1+AS03 group but not the H5N1 group significantly reduced production of most of the innate cytokines and chemokines that were diminished after vaccination with TIV (**Figure 5D**). Notably, we did not detect a change in the frequency or viability of classical monocytes and all of the detected cytokines were strongly induced by TLR stimulation (**Figure S6A, B, related to Figure 5**).

Next, we analyzed subjects using scATAC-seq and scRNA-seq. PBMC samples from 4 vaccinated individuals (2 H5N1, 2 H5N1+AS03) at days 0, 21, and 42 were enriched for DC subsets using flow cytometry and analyzed using droplet-based single-cell gene expression and chromatin accessibility profiling. After initial pre-processing, we obtained high quality chromatin accessibility data from 58,204 cells with an average of 2,745 uniquely accessible fragments which we used to generate an epigenomic map of the single immune cell landscape during H5N1 vaccination (**Figure 5E, DataS3**). In parallel, we used the scRNA-seq data to construct a gene expression map of the single immune cell landscape. We retained 11,213 high-quality transcriptomes with an average of 2,462 genes and 9,569 unique transcripts detected per cell and identified all major innate immune cell subsets (**Figure 5E, DataS3**). The different immune cell subsets were evenly distributed over all vaccine conditions and time points (**DataS3**).

Notably, using the scATAC-seq data, we observed a significant reduction in AP-1 accessibility in H5N1+AS03 but not H5N1 alone (**Figure 5F**). To further investigate the nature of the epigenomic changes after H5N1+AS03, we determined the differentially accessible regions at day 42 after vaccination compared to day 0 using a logistic regression model that corrects for library-size differences. Using overrepresentation analysis, we found that, similar to TIV, the predominantly negative DARs were enriched for TLR-, and cytokine-signaling pathways as well as innate immune activity (**Figure 5G**). Additionally, we observed a reduction in the expression of multiple AP-1 family members, including c-Fos and c-Jun as observed after vaccination with TIV (**Figure 5H**). Together, these findings suggest that vaccination with H5N1+AS03 induces epigenomic changes very similar to those observed after vaccination with TIV while vaccination with H5N1 alone only causes minor alterations to the epigenomic landscape.

### AS03 adjuvanted H5N1 influenza vaccine induces enhanced chromatin accessibility of the antiviral response loci

Despite the reduction in AP-1 accessibility, we observed an increase in chromatin accessibility at day 42 compared to day 0 for several TFs of the interferon-response factor (IRF) and STAT families (**Figure 6A**). These changes were observed in innate immune cell populations of subjects vaccinated with H5N1+AS03, but not with H5N1 alone. Further analysis of the kinetics revealed that these IRF- and STAT-related changes were already present after administration of the first vaccination at day 21 (pre-boost) (**Figure 6B**). Using the scRNA-seq dataset, we compared the expression of IRF and STAT family TFs before (day 0) and after prime (day 21) or boost (day 42) vaccination. We observed significant increases in the expression of IRF1 and STAT1 in multiple innate immune cell subsets after vaccination with H5N1+AS03, but not with H5N1 alone (**Figure 6C**). Notably, at a single-cell level, IRF accessibility was generally negatively correlated with AP-1 accessibility (**Figure 6D**), especially in dendritic cells. Next, we determined the log fold change in chromatin accessibility for peaks containing the IRF1 binding motif (**Figure 6E**). Indeed, we observed a significant change in accessibility in many peaks, most of which showed increased accessibility (**Figure 6E**). Importantly, amongst the genes with increased accessibility, we identified many interferon- and antiviral-related genes, including DDX58 (encoding the viral detector RIG-I), several interferon response genes (IFIT1, IFIT3, IFI30, ISG20, OASL), as well as the transcription factors IRF1 and IRF8. Enrichment analysis further demonstrated an enrichment of genes related to antiviral immunity (**Figure 6F**). IRF1, together with STAT1 and IRF8, orchestrates monocyte polarization in response to interferon gamma exposure **(Langlais et al., 2016);** IFN signaling, via JAK/TYK, leads to phosphorylation of IRF and STAT TFs **(Tamura et al., 2008)**. Indeed, we observed an increase in IFN gamma levels in plasma of vaccinated subjects immediately after prime and boost vaccination with H5N1+AS03, but not with H5N1 alone (**Figure 6G**). The levels of IP10, a cytokine that is produced by monocytes in response to IFN signaling, were also elevated (**Figure 6G**). This raises the possibility that IFN signaling could have induced the increased IRF accessibility.

**Figure 6.**
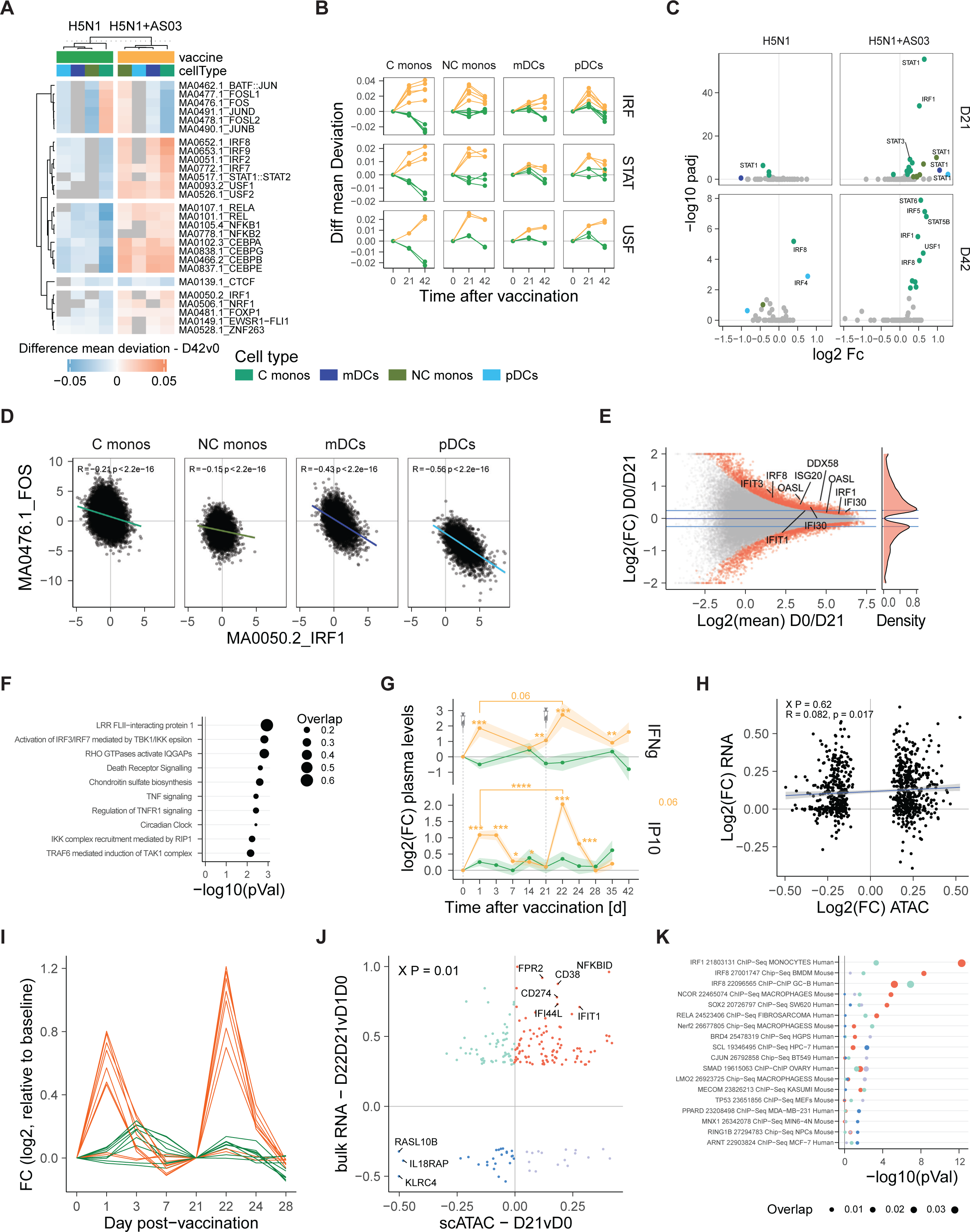
H5N1+AS03 induces epigenomic state of enhanced antiviral immunity. (A) Heatmap showing the change in chromatin accessibility at day 42 vs day 0 for the top5 transcription factors per subset. Color indicates the difference in accessibility, grey fields indicate non-significant changes (fdr > 0.05). (B) Line graph showing the difference in transcription factor (TF) accessibility during the course of the vaccine. (C) Volcano plot showing the change in gene expression for IRF/STAT TF genes. (E) MA plot showing the average accessibility and log2(FC) accessibility for genomic regions containing an IRF1 binding motif. Red color indicates regions with significantly changed accessibility (P <= 0.05). (F) Gene set enrichment analysis of significantly changed regions in E) occurring in at least 5% of C monos using the Reactome database. (G) Interferon alpha, gamma and IP10 levels were measured in plasma of vaccinated subjects at the indicated time points. Dots and lines indicate average, ribbons indicate standard error of mean. (H5N1/H5N1+AS03: IFNA, n = 5/11; IFNG, n = 7/14; IP10, n = 16/34) (H) Scatter plot showing the change in chromatin accessibility (x-axis) and the change in gene expression (y-axis) at day 21 vs day 0 for C monos (scATAC P <= 0.05 and occurring in at least 5% of cells). Indicated statistics are based on Pearson correlation analysis and Chi-square test. (I) Change in gene expression for selected antiviral and interferon-related BTMs in bulk RNA-seq analysis for subjects vaccinated with H1N1 (green) and H1N1+AS03 (orange) at indicated time points. (H1N1: n = 16, H1N1+AS03: n = 34. (J) Scatter plot showing the change in chromatin accessibility at day 21 vs day 0 in C monos (x-axis) and the significant change (p <= 0.05, log2(FC) > +/-0.03) in vaccine-induced gene expression at the booster vaccination compared to the prime vaccination (y-axis, Day22day21 vs Day1day0). Chi-square test was used to determine whether both variables were related. (K) Bubble plot showing enrichment results using the Encode TF target gene database. Color indicates the original of the analyzed genes in j).

Next, we determined whether the observed epigenomic changes were translated to changes in gene expression in resting monocytes. We assessed the relationship between the change in accessibility for significantly changed peaks carrying the IRF1 motifs and the change in gene expression for the same gene (**Figure 6H**). Notably, we detected a weak association between changes in accessibility and gene expression (Pearson correlation R = .082, p = .017, Chi-square p-value = 0.62) indicating that the increased accessibility has limited impact on homeostatic gene expression in these cells. Instead, we hypothesized that the changes in chromatin accessibility enhance the induced response to viral stimuli in activated cells. To test this hypothesis, we analyzed bulk RNA-seq data from 50 (16 H5N1, 34 H5N1+AS03) vaccinated subjects at time points before and after the prime (days 0, 1, 3, 7) and booster (days 21, 22, 24, 28) vaccination. As expected, antiviral- and interferon-related genes were upregulated at day 1 after each vaccination, especially in the group that received H5N1+AS03 (**Figure 6I**). Importantly, subjects receiving a H5N1+AS03 booster vaccination (day 22 vs day 21) displayed even higher levels of antiviral gene expression compared with the response to the prime vaccine (day 1 vs day 0) (**Figure 6I**). The booster vaccine was given at a time when the chromatin accessibility landscape of the innate immune system was altered suggesting that the increased accessibility in IRF loci might enable the enhanced response to the booster vaccine. To further test this hypothesis, we compared the increase in gene expression of antiviral- and interferon-related genes during booster compared to prime with the change in chromatin accessibility at day 21 compared to day 0 (**Figure 6J**). Indeed, we observed a significant association between both variables (Chi-square p-value = 0.01) and most genes with increased expression after booster vaccination also showed increased chromatin accessibility at the time the booster vaccine was administered. Genes with increased accessibility and enhanced expression were enriched for IRF1 transcription factor target genes (**Figure 6K**). In line with these observations, we also observed elevated levels of IP-10 and IFN gamma in plasma of individuals after the booster compared to prime vaccination (**Figure 6G**).

To determine if the observed epigenomic changes resulted in enhanced resistance to viral infections, we infected PBMCs at days 0, 21 and 42 with Dengue or Zika virus (**Figure 7A**). After infection, we cultured cells for 0, 24, and 48h and determined the viral copy number using qPCR (**Figure 7B**). We observed increased numbers of Zika and Dengue virus copies at 24h and reduction at 48h following the expected cycle of infection, replication, and eventual death of the host cells (**Figure 7C**). Next, we compared the viral titers at day 21 and 42 after vaccination with the pre-vaccination titers at day 0 for each subject. Strikingly, we observed a significant reduction in viral titers for both Dengue and Zika virus at day 21 after vaccination (**Figure 7D**). Importantly, in many subjects, we observed reduced viral titers as late as 42 days after initial vaccination (**Figure 7D**). Next, we determined the cytokine concentration in infected PBMC cultures at 24h after infection (**Figure 7E**). While Dengue and Zika virus induced the production of both IFNa and IFNg, we observed that Dengue virus suppressed the production of IP10 (**Figure 7E**). Finally, we correlated the change in viral titers at d0 compared to d21 with the change in vaccine-induced expression of antiviral genes that were associated with open chromatin (**Figure 6J** **red quadrant**). The majority of these genes correlated negatively with viral titers (**Figure 7F**). Strikingly, IRF1 was amongst the top genes negatively correlating (r < -0.8) with both Dengue and Zika titers (**Figure 7F**). Subjects with enhanced IRF1 expression at day 21 showed reduced viral titers at the same time point (**Figure 7G**). In addition, the antiviral gene ANKRD22, which is involved in immunity to both Dengue and Chikungunya infection **(Soares-Schanoski et al., 2019)**, was also highly negatively correlated with Zika and Dengue titers. Together, these data demonstrate that, despite the presence of AP-1-based suppression, AS03 induces an epigenomic state of enhanced antiviral immunity that enables increased production of interferons and enhanced control of heterologous viral infection.

**Figure 7.**
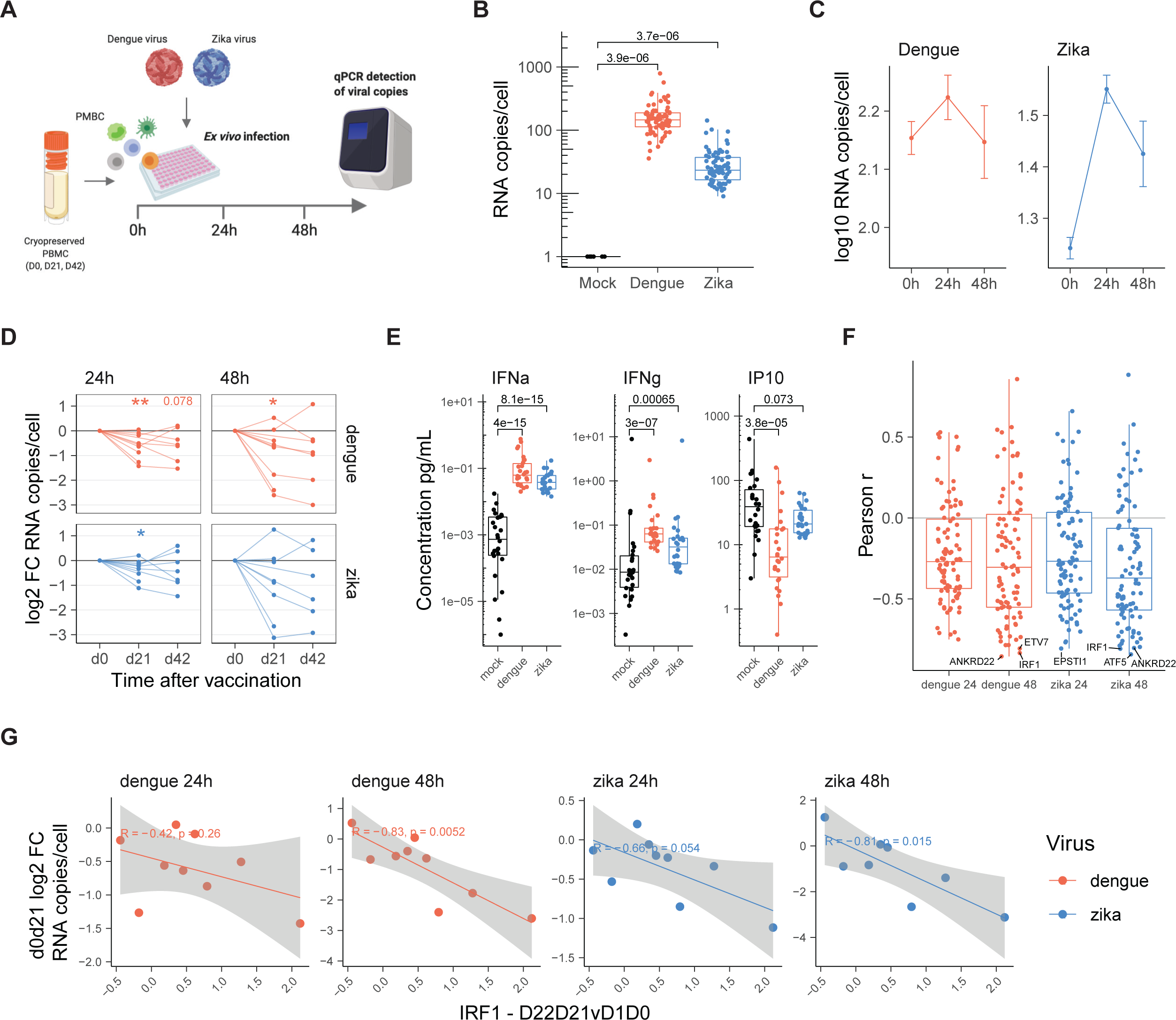
H1N1+AS03 induces enhanced resistance to in-vitro infection with heterologous viruses. (A) Schematic overview of the experiment. PBMCs from 10 healthy subjects at day 0, 21 and 42 after vaccination with H5N1+AS03 were infected with Dengue virus or Zika virus at an MOI of 1 and cultured for 0, 24 and 48 hours. After culture, viral copy numbers in cell pellet were determined via qPCR. (B) Boxplot showing viral titers in Dengue-, Zika-, and mock-infected samples. (C) Line graph showing the viral growth curve for Dengue virus (red) and zika virus (blue). Dots and lines indicate average, error bars indicate standard error of mean. n > 21 samples (D) Log2 fold change in viral titers relative to day 0 before vaccination. Wilcoxon signed rank test was used to compare changes within group; ** p <= 0.01, *p <= 0.05, n = 8-9. (E) Boxplot showing the concentration of IFNa, IFNg, and IP10 in Dengue-, Zika-, and mock-infected cultures at 24h after incubation. Wilcoxon rank-sum test was used to compare groups. (F, G) Pearson correlation analysis of the change in viral titers (d0 vs d21) with change in vaccine-induced, in-vivo expression of enhanced antiviral genes at prime (d0 vs d1) and boost (d21 vs d22) (red genes Figure 6g). F) Boxplot showing correlation coefficient per viral condition. G) Scatter plot showing change in vaccine-induced expression of IRF1 (x-axis) and viral titers (y-axis). B, E) Wilcoxon rank sum test was used to compare groups.

## Discussion

Here, we used several bulk- and single-cell approaches to construct the single-cell epigenomic landscape of immunity to three distinct influenza vaccines in humans: seasonal, as well as unadjuvanted and adjuvanted pre-pandemic influenza vaccines. Our results demonstrate that the seasonal inactivated influenza vaccine TIV and the adjuvanted pre-pandemic influenza vaccine (H5N1+AS03), induce profound and persistent global epigenomic changes in the peripheral immune system, especially in the myeloid compartment, and that these epigenomic changes are linked to alterations in the functionality of immune cells upon re-stimulation in-vitro and in-vivo. The observed changes were most pronounced at three to four weeks after vaccination but traces of an altered epigenomic landscape were still detectable as late as 180 days after initial vaccination.

Based on their molecular and functional characteristics, the observed epigenomic changes can be broadly classified into two distinct types: 1) a state of innate immune refractoriness that is characterized by reduced histone acetylation, reduced PADI4 levels, reduced AP-1 accessibility and diminished production of innate cytokines; 2) a state of heightened antiviral vigilance defined by increased IRF accessibility, elevated antiviral gene expression, increased interferon production, and, most importantly, enhanced control of heterologous viral infections. Importantly, both states can occur simultaneously and in the same single cell. While seemingly paradoxical, this superimposition might represent an evolutionary adaptation to avoid excess inflammatory host damage during late stages of infections, while maintaining a state of immunological vigilance against viral infections.

Our findings were unexpected as researchers previously observed that the live-attenuated BCG vaccine induces elevated H3K27ac levels in CD14^+^ monocytes which coincided with enhanced cytokine production in these cells **(Arts et al., 2018)**. In contrast, our results suggest that vaccine-induced epigenomic reprogramming of immune cells is more complex. Given the observed reduction in H3K27ac levels in association with immune refractoriness in this study, the possibility arises that histone acetylation could represent a bi-directional regulator, powered by epigenomically distinct states at the single-cell level, that can be raised or lowered to manipulate monocyte cytokine production accordingly, akin to a thermostat dial (**Figure S7**). In addition, our data demonstrate that multiple distinct epigenomic states, such as antiviral vigilance and immune refractoriness, can be superimposed within the same cell. This suggests a nuanced and compartmentalized reprogramming process that allows the independent adjustment of disparate chromatin loci to mediate distinct immunological processes in parallel. Importantly, this superimposition is encoded at the single-cell level as single monocytes and dendritic cells displayed elevated IRF and diminished AP-1 accessibility at the same time.

Single-cell analysis further revealed multiple clusters within the classical monocyte population based on differences in chromatin accessibility. Notably, all of these epigenomic subclusters existed before vaccination and their abundance within the pool of circulating cells shifted post vaccination driving the observed bulk level changes. The transcription factor families underlying the observed heterogeneity, AP-1 and CEBP, were previously described as key players in monocyte-to-macrophage differentiation **(Phanstiel et al., 2017)** and classical-to-non classical monocyte differentiation **(Guilliams et al., 2018)**, respectively. AP-1 signaling is also a central regulator of inflammation **(Bruggen et al., 1999; Das et al., 2009; Fontana et al., 2015; Fujioka et al., 2004; Hannemann et al., 2017; Ventura et al., 2003)** and our Hotspot analysis revealed differences in accessibility at inflammatory loci between epigenomic subclusters. This might suggest that distinct functional and ontogenetic fates could be imprinted within the epigenome of single monocytes. Indeed, it was recently hypothesized that classical monocytes could represent a heterologous population of cells, some pre-committed to tissue infiltration and macrophage differentiation and others primed for differentiation into non-classical monocytes **(Guilliams et al., 2018)**. Two conceptual questions arise from these findings: 1) Do the observed epigenomic subclusters represent stable, distinct epigenomic states or merely degrees on an epigenomic spectrum? 2) Are circulating monocytes able to transition from one state to another in response to vaccination, or does vaccination influence the commitment decisions already at the progenitor level in the bone marrow?

Our systems biology analysis also extends the current CD14^+^ monocyte-focused understanding of vaccine-induced epigenomic reprogramming with insights into other circulating cells of the innate immune system. Previously, it was not known whether CD14^-^ monocytes or dendritic cells would exhibit epigenomic changes after vaccination in humans. Here, we show that influenza vaccination also induces lasting epigenomic changes in mDCs that share many of the molecular characteristics with those observed in classical monocytes. In contrast, non-classical CD14^-^CD16^+^ monocytes and pDCs presented less pronounced and more short-lived alterations in their epigenomic state. The molecular events that enable this selectivity are unknown and its discovery came as a surprise as circulating non-classical monocytes are thought to directly differentiate from classical monocytes **(Patel et al., 2017)**. However, in light of the observed epigenomic subclusters within the monocyte pool, it is conceivable that the vaccine-induced epigenomic changes are not affecting classical monocytes primed for differentiation into non-classical monocytes. A similar mechanism is conceivable for pDCs, which are thought to branch off early from mDC progenitors during development **(See et al., 2017)**.

With respect to the molecular mechanisms driving the epigenomic changes, we observed that the state of antiviral vigilance was associated with enhanced IRF1 and STAT1 activity. It is established that IRF and STAT signaling promotes antiviral immunity **(Tamura et al., 2008)** and KO models lacking IRF1 or STAT1 are more susceptible to viral infection **(Meraz et al., 1996; Panda et al., 2019)**. In contrast, the state of immune refractoriness was associated with a global reduction in histone acetylation and chromatin accessibility. The magnitude of the observed changes suggests a comprehensive switch towards a broadly restrictive chromatin state **(Allis and Jenuwein, 2016)**. Our TF motif-based analysis revealed that especially AP-1 loci are affected by this process. AP-1 is a dimeric TF composed of different members of the FOS, JUN, ATF, and JDP families and our gene expression analysis suggests that multiple members including FOS, JUN, JUNB, and ATF3 are involved. While the role of AP-1 as a key regulator of differentiation, inflammation and polarization in myeloid cells is well described **(Behre et al., 1999; Bruggen et al., 1999; Das et al., 2009; Fontana et al., 2015; Fujioka et al., 2004; Hannemann et al., 2017; Monick et al., 1999; Phanstiel et al., 2017; Tsai et al., 2000; Ventura et al., 2003)**, recent research also positions it as a central epigenomic regulator **(Arias et al., 1994; Beisaw Arica et al., 2020; Biddie et al., 2011; Phanstiel et al., 2017; Zanger et al., 2001)**.

A fundamental question in the context of our results concerns the mechanisms by which vaccination induces such long-lasting epigenetic changes in myeloid cells. The half-lives of most DC and monocyte subsets are known to be only a few days **(van Furth and Cohn, 1968; Kamath et al., 2000)**. Therefore, it is unclear how epigenetic changes acquired by a DC or monocyte responding to a vaccine might be maintained for several weeks or months. Multiple explanations are conceivable: for instance, the phenomenon of innate memory could simply be caused by the effects of an ongoing adaptive immune response on innate immune cells (via paracrine signaling of cytokines such as interferon-gamma), rather than being an intrinsic property of innate immune cells. Furthermore, innate memory could be maintained by some long-lived population of innate immune cells, like memory T and B cells, and such cells could respond with enhanced vigor to a secondary vaccination or infection. Finally, it is possible that epigenetically reprogrammed myeloid cells in the periphery are continually replenished by altered myeloid cell precursors in the bone marrow **(Cirovic et al., 2020; Kaufmann et al., 2018; Mitroulis et al., 2018)**.

From a biological perspective, we made multiple observations that suggest a direct link between the epigenomic state and immune protection. Our results from the H5N1+AS03 vaccine revealed that PBMCs from vaccinated individuals control infection with the heterologous Dengue and Zika virus more efficiently than pre-vaccination PBMCs. These results, in combination with the enhanced expression of antiviral genes and increased levels of IP-10 and IFN gamma production in-vivo, suggest that the epigenomic state of antiviral vigilance might provide non-specific protection against viral infections unrelated to the vaccine virus. Elevated levels of IFN gamma production at day 1 after booster vaccination were also detected with AS03-adjuvanted vaccines in the context of Hepatitis **(Burny et al., 2017)** suggesting that antiviral vigilance might also be induced by other vaccines containing AS03. In contrast, TIV induced a profound state of immune refractoriness at four weeks after vaccination. It is important to highlight that there is ample evidence that TIV does prevent influenza **(Centers for Disease Control and Prevention, 2020)** and our own study found induction of robust anti-influenza antibody titers **(Hagan et al., 2019)**. Given the observed immune refractoriness, it could be beneficial to administer TIV together with an adjuvant, such as AS03, especially in vulnerable populations. This adjuvanted TIV would overcome the induced immune refractoriness with an epigenomics-driven state of antiviral vigilance. Indeed, a phase 3 clinical trial comparing the response to TIV vs TIV+AS03 in more than 43,000 Elderly individuals demonstrated that TIV+AS03 led to a profound reduction in all-cause death and pneumonia compared to TIV alone while influenza-specific immunity was only somewhat increased **(McElhaney et al., 2013)**. It is worth mentioning that an inactivated seasonal influenza vaccine with the adjuvant MF59 is currently licensed for the use in individuals older than 65 years. Like AS03, MF59 is based on squalene. While subsequent double-blinded studies comparing adjuvanted vs. non-adjuvanted influenza vaccines are needed to definitively prove the clinical benefit, our results demonstrate a previously unknown mechanism of action for adjuvants to provide non-specific protection via epigenomic reprogramming.

In conclusion, we combined multiple single-cell high-throughput techniques to investigate how vaccination with three distinct influenza vaccines alters the epigenomic immune cell landscape at the single-cell level. Our results demonstrate persistent epigenomic changes in myeloid cells after influenza vaccination which were linked to altered innate immune cell functionality and heterologous protection against non-vaccine viruses. Single-cell analysis revealed the presence of epigenomically distinct subcluster within the monocyte population. Our findings have implications for the design of future vaccines and might inspire the development of epigenomic adjuvants that provide broad, non-specific protection by manipulating the epigenomic landscape.

## Data Availability

EpiTOF data is available at: will be available upon publication. Bulk ATAC-seq and RNA-seq data is available here: will be available upon publication. ScATAC-seq and scRNA-seq data are deposited at: will be available upon publication.

## Acknowledgements

We are grateful to Mary Bower and the Hope Clinic staff and faculty who conducted the clinical work. We thank the Human Immune Monitoring Center (HIMC), the Parker Institute for Cancer Immunotherapy (PICI), and the Stanford FACS facility for maintenance and access to flow cytometers and FACS sorting. We particularly acknowledge Mary Rieck for extraordinary support with FACS sorting. We thank the HIMC for assistance with Luminex analysis, especially Yael Rosenberg-Hasson. We are thankful for the Stanford Functional Genomics Facility for technical assistance; Dhananjay Wagh and Ed Kim for library preparation, John Coller for data analysis. We are grateful to Fabian Mueller, Arwa Kathiria and Will Greenleaf for assisting during the early stages of the ATAC-seq analysis. We thank ActiveMotif for their customized ATAC-seq services, especially Chris Balagtas and Paul Labhart. Next generation sequencing services were provided by the Yerkes NHP Genomics Core which is supported in part by NIH P51 OD011132. Sequencing data was acquired on an Illumina NovaSeq6000 funded by NIH S10 OD026799. IP-10 plasma Luminex analysis was performed in the Immunology Unit of the Regional Biocontainment Laboratory at Duke, which received partial support for construction from the National Institutes of Health, National Institute of Allergy and Infectious Diseases (UC6-AI058607). Cartoons were created with BioRender.com. We thank Philippe Boutet, Nathalie Arts, Olivia Furstoss, Vanesa Bol, and Margherita Coccia for critically reviewing the manuscript.

## Materials & Methods

### RESOURCE AVAILABILITY

#### Lead Contact

Further information and requests for resources and reagents should be directed to and will be fulfilled by the lead contact, Bali Pulendran.

#### Materials Availability

This study did not generate new unique reagents.

### EXPERIMENTAL SUBJECT DETAILS TIV

The study design was as described in phase 1 of the original publication **(Hagan et al., 2019)** and the study was conducted in Atlanta, GA. In brief, during the 2014-2015 seasons, we enrolled a total of 21 healthy adults who were randomized into antibiotics-treated (n = 10) and control (n = 11) groups. Subjects were males and non-pregnant females between the ages of 18-40 who met the eligibility criteria as listed on clinicaltrials.gov (NCT02154061). Subject demographics are listed in (**DataS1**). The antibiotics treatment consisted of a cocktail of neomycin, vancomycin, and metronidazole, all given orally, for five days. Antibiotic treatment started 3 days before the day of vaccination and continued until one day after for the antibiotics-treated group. All the study participants were vaccinated with Sanofi Pasteur’s TIV vaccine, Fluzone, for the 2014-2015 season **(DataS1)**. Written informed consent was obtained from each subject and protocols were approved by Institutional Review Boards of Emory University.

#### H5N1/H5N1+AS03

This study was conducted in Atlanta, GA. Subjects were males and non-pregnant females who met the eligibility criteria as listed on clinicaltrials.gov (NCT01910519). We enrolled a total of 50 healthy adults who were randomized into two groups receiving either the adjuvanted (H5N1+AS03, n=34) or unadjuvanted (H5N1, n=16) GSK avian influenza vaccine. While both vaccines contained split-virion (A/Indonesia/5/2005) inactivated hemagglutinin antigen, the adjuvanted vaccine additionally contained the AS03 adjuvant system (containing DL-alpha-tocopherol and squalene in an oil-in-water emulsion). Subject demographics are listed in (**DataS1**). Written informed consent was obtained from each subject and protocols were approved by Institutional Review Boards of Emory University.

#### In-vitro stimulation and intracellular flow cytometry experiments

Samples from healthy subjects were collected at Stanford Blood Center or derived from the before-vaccination time point of a previous vaccination trial **(Nakaya et al., 2015)**. All subjects provided a confidential medical history card and completed informed consent to donate blood for clinical or research uses. We exclude subjects with known diseases, including but not limited to HIV, and hepatitis infections. Purification of buffy coat or LRS chamber from whole blood was performed at Stanford Blood Center to enrich for leukocytes prior to PBMC isolation.

From the vaccination trial (Nakaya et al., 2015), only samples from subjects aged 26 – 41 were selected for this paper. Samples were only selected from the before vaccination time point at day 0. Written informed consent was obtained from each subject with institutional review and approval from the Emory University Institutional Review Board.

### METHOD DETAILS

#### Cells, plasma and RNA isolation

Peripheral blood mononuclear cells (PBMCs) and plasma were isolated from fresh blood (CPTs; Vacutainer with Sodium Citrate; BD), following the manufacturer’s protocol. For samples from Stanford Blood Center, PBMCs isolated from whole blood, buffy coat or LRS chamber by Ficoll density gradient centrifugation using Ficoll-Paque PLUS (GE Healthcare, #17-1440-02). PBMCs were frozen in DMSO with 10% FBS and stored at –80C and then transferred on the next day to liquid nitrogen freezers (–196C). Plasma samples from CPTs were stored at –80C. Trizol (Invitrogen) was used to lyse fresh PBMCs (1 mL of Trizol to ∼1.5×10^6 cells) and to protect RNA from degradation. Trizol samples were stored at –80C.

#### Mass cytometry sample processing, staining, barcoding and data acquisition

Cryopreserved PBMCs were thawed and incubated in RPMI 1640 media (ThermoFisher) containing 10% FBS (ATCC) at 37°C for 1 hour prior to processing. Cisplatin (ENZO Life Sciences) was added to 10 µM final concentration for viability staining for 5 minutes before quenching with CyTOF Buffer (PBS (ThermoFisher) with 1% BSA (Sigma), 2mM EDTA (Fisher), 0.05% sodium azide). Cells were centrifuged at 400g for 8 minutes and stained with lanthanide-labeled antibodies (**DataS2**) against immunophenotypic markers in CyTOF buffer containing Fc receptor blocker (BioLegend) for 30 minutes at room temperature (RT). Following extracellular marker staining, cells were washed 3 times with CyTOF buffer and fixed in 1.6% PFA (Electron Microscopy Sciences) at 1×106 cells/ml for 15 minutes at RT. Cells were centrifuged at 600g for 5 minutes post-fixation and permeabilized with 1 ml ice-cold methanol (Fisher Scientific) for 20 minutes at 4°C. 4 ml of CyTOF buffer was added to stop permeabilization followed by 2 PBS washes. Mass-tag sample barcoding was performed following the manufacturer’s protocol (Fluidigm). Individual samples were then combined and stained with intracellular antibodies in CyTOF buffer containing Fc receptor blocker (BioLegend) overnight at 4°C. The following day, cells were washed twice in CyTOF buffer and stained with 250 nM 191/193Ir DNA intercalator (Fluidigm) in PBS with 1.6% PFA for 30 minutes at RT. Cells were washed twice with CyTOF buffer and once with double-deionized water (ddH2O) (ThermoFisher) followed by filtering through 35μm strainer to remove aggregates. Cells were resuspended in ddH2O containing four element calibration beads (Fluidigm) and analyzed on CyTOF2 (Fluidigm).

#### Bulk stimulation experiment

Aliquots of thawed PBMCs from the EpiTOF experiment described above were washed and resuspended in RPMI 1640 (Corning, 10-040-CV) containing 10% FBS (Corning, 35-011-CV) and 1x Antibiotics/Antimycotics (Lonza, 17-602E) [complete media abx] at 4×10^6 cells/mL. 100 µL of cell solution were added to each well of a 96-well round-bottomed tissue culture plate and mixed with 100 µL of either complete media abx (unstim), a cocktail of synthetic TLR ligands mimicking bacterial pathogens (bac: 0.025 µg/mL LPS, 0.3 µg/mL Flagellin, 10 µg/mL Pam3CSK4), or a cocktail of synthetic TLR ligands mimicking viral pathogens (vir: 4 µg/mL R848, 25 µg/mL pI:C). Depending on cell numbers, PBMCs from each sample were stimulated with all 3 conditions in duplicate. After 24h of incubation at 37C and 5% CO2, cells were spun down, supernatant was carefully transferred into new plates, and immediately frozen at -80C until further analysis using Luminex.

#### Luminex TIV

The Luminex assay was performed by the Human Immune Monitoring Center, Stanford University School of Medicine. Human 62-plex custom Procarta Plex Kits (Thermo Fisher Scientific) were used according to the manufacturer’s recommendations with modifications as follows: Briefly, Antibody-linked magnetic microbeads were added to a 96-well plate along with custom Assay Control microbeads (Assay Chex) by Radix Biosolutions. The plates were washed in a BioTek ELx405 magnetic washer (BioTek Instruments). Neat Cell culture supernatants (25ul) and assay buffer (25ul) were added to the 96 well plate containing the Antibody-coupled magnetic microbeads, and incubated at room temperature for 1 h, followed by overnight incubation at 4°C. Room temperature and 4°C incubation steps were performed on an orbital shaker at 500–600 rpm. Following the overnight incubation, plates were washed in a BioTek ELx405 washer (BioTek Instruments) and then kit-supplied biotinylated detection Ab mix was added and incubated for 60 min at room temperature. Each plate was washed as above, and kit-supplied streptavidin–PE was added. After incubation for 30 min at room temperature, wash was performed as described, and kit Reading Buffer was added to the wells. Each sample was measured in two technical replicates where cell numbers allowed. Plates were read using a FlexMap 3D Instrument (Luminex Corporation). Wells with a bead count <50 were flagged, and data with a bead count <20 were excluded.

#### Luminex H5N1/H5N1+AS03

This assay was performed by the Human Immune Monitoring Center at Stanford University. A custom 41 plex from EMD Millipore kits was assembled and included: 1. A Pre-mixed 38 plex Milliplex Human Cytokine/Chemokine kit (CAT# HCYTMAG-60K-PX38) 2. ENA78/CXCL5 (CAT# HCYP2MAG-62K-01) 3. IL-22 (CAT# HTH17MAG-14K-01). 4. IL-18 (HIL18MAG-66K). Manufacturer’s recommendations were followed with modifications described. Briefly: neat supernatant samples (25ul) were mixed with antibody-linked magnetic beads in a 96-well plate containing assay buffer, for an overnight incubation at 4°C. Cold and Room temperature incubation steps were performed on an orbital shaker at 500-600 rpm. Plates were washed twice with wash buffer in a BioTek ELx405 washer (BioTek Instruments). Following one-hour incubation at room temperature with biotinylated detection antibody, streptavidin-PE was added for 30 minutes. Plates were washed as above, and PBS was added to wells for reading in the Luminex FlexMap3D Instrument with a lower bound of 50 beads per sample per cytokine. Each sample was measured in duplicate wells where cell numbers allowed. Custom Assay Chex control beads were added to all well (Radix Biosolutions). Wells with a bead count <50 were flagged, and data with a bead count <20 were excluded.

#### H3K27ac antibody conjugation

*α*-H3K27ac antibody was labeled using the Lightning-Link Rapid DyLight 488 Antibody Labeling Kit according to manufacturer’s instructions (Novus Biologicals, 322-0010). In brief, 100 µg of antibody was mixed with 10 µL of LL-Rapid modifier reagent and added onto the lyophilized dye. After mixing, solution was incubated at room temperature overnight in the dark. The next morning, 10 µL of LL-Rapid quencher reagent was added.

#### In-vitro stimulation and intracellular flow cytometry experiments

Cryopreserved PBMCs were thawed, counted, and resuspended in RPMI 1640 (Corning, 10-040-CV) supplemented with 10% FBS (Corning, 35-011-CV) [complete media] at a concentration of 4×10^6 cells/mL. Next, 150µL of cell suspension (6×10^5 cells) was added to each well of a 96-well round-bottomed tissue culture plate and mixed with 50 µL of inhibitor solution containing either Trichostatin A (TSA; CST, 9950S), A-485 (Tocris, 6387), or Cl-Amidine (EMD Millipore, 506282) in complete media. After 2h of incubation at 37C and 5% CO2, the cells were stimulated by adding either LPS (0.025 µg/mL; Invivogen, tlrl-pb5lps) or R848 (4 µg/mL; Enzo Life Sciences, ALX-420-038-M005) to the cultures. After another 2h of incubation, Brefeldin A (10 µg/mL; Sigma Aldrich, B7651-5MG) was added to all cultures and cells were incubated for a final 4h.nAfter a total of 8h of incubation, cells were washed twice with 150 µL PBS (GE Life Sciences, SH30256.LS) and stained for viability using 100 µL of Zombie UV Fixable Viability Dye in PBS (1:1000; Biolegend, 423108). After incubating for 30 minutes at 4C in the dark, cells were washed twice with 150 µL PBS and blocked with 100 µL of PBS supplemented with 5% FBS, EDTA (2 mM; Corning, 46-034-cl), and human IgG (5 mg/mL; Sigma Aldrich, G4386-5G) [blocking buffer] for 15 minutes at 4C in the dark. After incubation, cells were stained for surface markers with 100 µL of antibody cocktail containing *α*-CD14 BUV805, *α*-CD3, CD19, CD20 BUV737, *α*-CD123 BUV395, *α*-HLA-DR BV785, *α*-CD16 BV605, *α*-CD56 PE-CY7, *α*-CD11c APC-eFluor780 in blocking buffer for 20 minutes at 4C in the dark. Next, cells were washed twice with 150 µL PBS, and fixed in 200 µL eBioscience Foxp3 Fixation/Permeabilization solution (ThermoFisher Scientific, 00-5523-00) for 30 minutes at 4C in the dark. Afterwards, cells were washed twice with 100 µL eBioscience Foxp3 permeabilization buffer and blocked with 100 µL permeabilization buffer containing human IgG (5 mg/mL) overnight at 4C in the dark. Cells were washed and stained for intracellular markers with 25 µL of antibody cocktail containing *α*-IL-1b Pacific Blue, *α*-H3K27ac DyLight 488, *α*-TNFa PE-Dazzle, *α*-p-c-Jun PE, and *α*-H3 AF647 in permeabilization buffer containing human IgG (5 mg/mL) for 60 minutes at 4C in the dark. Finally, cells were washed twice with 150 µL of permeabilization buffer, resuspended in 100 µL PBS containing 0.5% FBS and 2 mM EDTA [FACS buffer], and acquired using a BD FACSymphony flow cytometer. Data was analyzed using Flowjo X software (BD). Briefly, cells were identified via FSC/SSC, doublets were discarded via SSC-A/SSC-H and FSC-A/FSC-H gates, and dead cells were discarded as Zombie UV Fixable Viability Dye high. Monocytes were then identified as CD3^-^CD19^-^CD20^-^ and CD14^+^.

#### FACS sorting – bulk ATAC-seq/RNA-seq

Cryopreserved PBMCs were thawed, washed, counted, and resuspended in PBS (GE Life Sciences, SH30256.LS). 5-10×10^6 cells were washed once more with 2 mL of PBS and stained for viability using 500 µL of Zombie UV Fixable Viability Dye in PBS (1:1000; Biolegend, 423108). After incubating for 30 minutes at 4C in the dark, cells were washed with 2 mL of PBS and resuspended in 500 µL blocking buffer. After spinning cells down, supernatant was discarded, and cells were resuspended in 50 µL antibody cocktail containing *α*-CD3, CD19, CD20 BUV737, *α*-CD123 BUV395, *α*-HLA-DR BV785, *α*-CD14 BV605, *α*-CD56 BV510, *α*-CD1c BV421, *α*-CD327 AF488, *α*-CD370 PE, *α*-CD11c APC-eFluor780, *α*-CD15 AF700, and *α*-Axl APC in blocking buffer. Cells were stained for 15 minutes at 4C in the dark. Finally, cells were washed with 2 mL of FACS buffer, resuspended in PBS containing 5% FBS at 10-20×10^6 cells/mL, and stored at 4C before sorting on a FACS Aria Fusion (BD). During sort, live innate cells were identified by gating on Viability Dye^-^ CD3^-^CD19^-^CD20^-^ cells. Within this population, CD14^+^ monocytes were identified as CD14^+^, mDCs were identified as CD14^-^ CD56^-^HLA-DR^+^CD16^-^CD11c^+^CD123^-^, and pDCs were identified as CD14^-^CD56^-^HLA-DR^+^CD16^-^CD11c^-^CD123^+^.

#### Omni ATAC-seq of purified immune cells

Atac was performed on purified innate immune cell subsets immediately after sorting based on the low-input Omni-Atac protocol described before **(Corces et al., 2017)**. In brief, 1,500 – 5,500 cells were washed with ATAC resuspension buffer (10 mM Tris-HCl pH 7.5 [Invitrogen, 15567027], 10 mM NaCl [Invitrogen, AM9760G], 3 mM MgCl2 [Invitrogen, AM9530G], in water [Invitrogen, 10977015]) and supernatant was carefully aspirated, first using a P1000, then a P200 pipette. Next, 10 µL transposition mix (0.5 µL Tn5, 0.1 µL 10% Tween-20, 0.1 µL 1% Digitonin, 3.3 µL PBS, 1 µL water, and 5 µL tagmentation buffer) was added to the pellet and cells were resuspended by pipetting up and down 6 times. Tagmentation buffer was prepared locally by resuspending 20 mM Tris-HCl pH 7.5, 10 mM MgCl2, and 20% Dimethyl Formamide (Sigma Aldrich, D4551-250ML) in water. Cells were incubated at 37C for 30 minutes under constant mixing. After tagmentation, the reaction was cleaned up using the MinElute PCR Purification Kit (Qiagen, 28006) according to manufacturer’s instructions. Cleaned DNA was eluted in 21 µL of elution buffer, stored at -20C, and shipped to Active Motif for sequencing library preparation. At Active Motif, tagmented DNA was amplified with 10 cycles of PCR using customized Nextera PCR Primers 1 and 2 (see Key Resource table), and purified using Agencourt AMPure SPRI beads (Beckman Coulter, A63882). Resulting material was quantified using the KAPA Library Quantification Kit for Illumina platforms (Roche, 07960255001), and sequenced with PE42 sequencing on the NextSeq 500 sequencer (Illumina).

#### Bulk RNA-seq of purified immune cells

Bulk RNA-seq was performed on purified CD14^+^ monocytes after sorting. In brief, after sorting, 5,500 cells were washed, resuspended in 350 µL chilled Buffer RLT (Qiagen, 79216) supplemented with 1% beta-Mercaptoethanol (Sigma, M3148-25ML), vortexed for 1 minute, and immediately frozen at -80C. RNA was isolated using the RNeasy Micro kit (Qiagen, 74004) with on-column DNase digestion. RNA quality was assessed using an Agilent Bioanalyzer and total RNA was used as input for cDNA synthesis using the Clontech SMART-Seq v4 Ultra Low Input RNA kit (Takara Bio, 634894) according to the manufacturer’s instructions. Amplified cDNA was fragmented and appended with dual-indexed bar codes using the NexteraXT DNA Library Preparation kit (Illumina, FC-131-1096). Libraries were validated by capillary electrophoresis on an Agilent 4200 TapeStation, pooled at equimolar concentrations, and sequenced on an Illumina NovaSeq6000 at 100SR, yielding 20-25 million reads per sample.

#### FACS sorting – scATAC-seq/RNA-seq

Cryopreserved PBMCs were thawed and innate immune cell subsets were isolated using FACS as described above (**FACS sorting – bulk ATAC-seq/RNA-seq**). Within the live gated cells, CD14^+^ monocytes were identified as CD14^+^ (fraction A) while a mixture of the remaining monocyte and dendritic cell subsets was identified as CD14^-^ CD56^-^HLA-DR^+^ (fraction B). After sorting and depending on the number of isolated cells, fraction A and B were mixed at a 2:1 ratio to yield a solution of monocytes and dendritic cells enriched for CD14^-^ cells.

#### scRNA-seq

FACS-purified cells were resuspended in PBS supplemented with 1% BSA (Miltenyi), and 0.5 U/μL RNase Inhibitor (Sigma Aldrich). About 9,000 cells were targeted for each experiment. Cells were mixed with the reverse transcription mix and subjected to partitioning along with the Chromium gel-beads using the 10X Chromium system to generate the Gel-Bead in Emulsions (GEMs) using the 3’ V3 chemistry (10X Genomics). The RT reaction was conducted in the C1000 touch PCR instrument (BioRad). Barcoded cDNA was extracted from the GEMs by Post-GEM RT-cleanup and amplified for 12 cycles. Amplified cDNA was subjected to 0.6x SPRI beads cleanup (Beckman, B23318). 25% of the amplified cDNA was subjected to enzymatic fragmentation, end-repair, A tailing, adapter ligation and 10X specific sample indexing as per manufacturer’s protocol. Libraries were quantified using Bioanalyzer (Agilent) analysis. Libraries were pooled and sequenced on an NovaSeq 6000 instrument (Illumina) using the recommended sequencing read lengths of 28 bp (Read 1), 8 bp (i7 Index Read), and 91 bp (Read 2).

#### scATAC-seq

FACS-purified cells were processed for single nuclei ATAC-seq according to the manufacturer’s instructions (10x Genomics, CG000168 Rev D). Briefly, nuclei were obtained by incubating PBMCs for 3.20 minutes in freshly prepared Lysis buffer following manufacturer’s instructions for Low Cell Input Nuclei Isolation (10x Genomics, CG000169 Rev C). Nuclei were washed and resuspended in chilled diluted nuclei buffer (10x Genomics, 2000153). Next, nuclei were subjected to transposition for 1h at 37C on the C1000 touch PCR instrument (BioRad) prior to single nucleus capture on the 10x Chromium instrument. Samples were subjected to post GEM cleanup, sample index PCR, cleanup and library QC prior to sequencing according to the protocol. Samples were pooled, quantified and sequenced on NovaSeq 6000 instrument (Illumina) with at least minimum recommended read depth (25000 read pairs/nucleus).

#### IFNa SIMOA

Frozen plasma was shipped to Qunaterix and analyzed using the Simoa® IFN-α Advantage Kit (Quanterix, 100860) according to manufacturer’s instructions. In brief, plasma and reagents were thawed at room temperature. Cailbrators, controls, and plasma were transferred to assay plates. Beads were vortexed for 30 seconds and prepared reagents and samples were loaded into a HD-1/HD-X instrument and analyzed with standard settings. All samples were run in duplicate.

#### Detection of IFNa and IFNg in plasma and cell culture supernatants

Frozen plasma or supernatant was thawed at room temperature and analyzed using the IFNa and IFNg Human ProQuantum Immunoassay Kits according to manufacturer’s instructions. In brief, samples were mixed with assay dilution buffer at a 1:5 or 1:2 ratio and protein standard was serially diluted in assay dilution buffer. Next, Antibody-conjugates A and B were mixed with Antibody-conjugate dilution buffer and added to each well of a 96-well qPCR plate (Bio-Rad, #HSP9601). Next, diluted sample or standard were added to each well and mixtures were incubated for 1h at room temperature in the dark. Finally, Master max and Ligase were mixed and added to each well. QPCR was conducted on a CFX96 Touch Real-Time Detection System (Biorad) using the recommended instrument settings. After measurements were completed, CT values were calculated using a regression model and exported to the ProQuantum Cloud app that accompanied the kit (apps.thermofisher.com/apps/proquantum). ProQuantum Cloud app was then used to construct a standard curve and calculate protein concentrations from CT values.

#### IP-10 plasma Luminex

Plasma biomarker levels were assayed using a 10-analyte multiplex bead array (fractalkine, IL-12P40, IL-13, IL-1RA, IL-1b, IL-6, IP-10, MCP-1, MIP-1α, TNFβ; Millipore) prepared according to the manufacturer’s recommended protocol and read using a Bio-Plex 200 suspension array reader (Bio-Rad). Data were analyzed using Bio-Plex manager software (Bio-Rad).

#### Viral infection assay

Dengue virus (DENV-2, Strain Thailand/16681/84) and Zika virus (PRVABC59) were propagated and titrated on Vero cells and stored at -80C until infection. Cryopreserved human PBMCs were thawed, washed, counted, and resuspended in RPMI 1640 (Thermo Fisher, 72400-047) supplemented with 10% FBS (Corning, 35-011-CV), 1mM Sodium pyruvate (Lonza, 13-115E), and 1x Penicillin/Streptomycin (Lonza, 17-602E) at 1.5×10^6 cells/mL. 200 µL of cell solution (3×10^5 cells) was added to each well of a 96-well round-bottomed tissue culture plate and cells were rested in plates for 4h at 37C and 5% CO_2_. After resting, PBMCs were infected with DENV-2 or ZIKV at MOI 1. At 0h, 24h, 48h post infection, PBMCs and supernatant were collected for RNA purification and cytokine analysis, respectively. Supernatants were immediately frozen at -20C and stored until analysis. Cells were suspended in RNA lysis buffer and kept at -20C until analysis. RNA was purified using the Purelink RNA kit according to manufacturer recommendations (Thermo Fisher Scientific, #12183052). For viral load detection, quantitative reverse transcription PCR (qRT-PCR) was conducted using Luna universal probe one-step RT-PCR kit (NEB, #E3006) on a CFX96 C1000 Touch Real-Time Detection System with 96-well plates (Bio-Rad, #HSP9601). RNA standards (ATCC, # VR-3229SD, VR-1843DQ) were used to generate standard curves. Viral RNA copies were normalized by cell number. Utilized primers and probes are listed in the Key Resources table.

#### Detection of IP-10 in culture supernatant

Culture supernatants were thawed at room temperature and analyzed using the IP-10 enzyme-linked immunosorbent assay (R&D Systems, DIP100) according to the manufacturer’s instructions. In brief, samples were thawed at room temperature and mixed with assay dilution buffer at 1:2 ratio. Protein standard was serially diluted in assay dilution buffer. Samples and standards were incubated in plate for 2h at room temperature. Plates were washed and then incubated with human IP-10 conjugate for 2h at room temperature. After wash, substrate solution was added for 30min. Finally, stop solution was added, A450 and A595 were read on a plate reader (Bio-Rad, iMARK). The concentration of IP-10 was determined by the number of A450-A595 based on the standard curve.

### QUANTIFICATION AND STATISTICAL ANALYSIS

**Immune Cell Population Definitions and EpiTOF Data Pre-Processing**

Raw data were pre-processed using FlowJo (FlowJo, LLC) to identify cell events from individual samples by palladium-based mass tags, and to segregate specific immune cell populations by immunophenotypic markers. A detailed gating hierarchy is described in **DataS2** (TIV & H5N1/H5N1+AS03). Single-cell data for various immune cell subtypes from individual subjects were exported from FlowJo for downstream computational analyses.

#### EpiTOF analysis

The exported Flowjo data were then normalized following the approach described in **(Cheung et al., 2018)**. In brief, the value of each histone mark was regressed against the total amount of histones, represented by measured values of H3 and H4. For sample level analyses, the values of each histone mark were averaged for each cell type in each sample. Distances of HSC from lymphoid and myeloid epigenetic profiles were obtained by first computing centers of the epigenetic profiles for the two lineages, and then computing Euclidean distances from the centers for each individual HSC. Distances of HSC from epigenetic profiles of specific cell types were similarly obtained by computing Euclidean distances from the centers of the epigenetic profiles for each cell type. Statistical significance of the differences between groups at the sample level was assessed by computing an effect size with Hedges’g formula **(Hedges and Olkin, 2014)**. All p-values were corrected for multiple comparisons with the Benjamini-Hochberg method **(Benjamini and Hochberg, 1995)**. Dimensionality reduction was performed with applying UMAP **(McInnes et al., 2020)**. For single cell analyses, the normalized values were used as input. Correlation between variables was computed using Pearson’s correlation coefficient. All the analyses were performed using the R framework for statistical computing (Version 3.6.3) **(R Core Team, 2020)**.

#### TIV bulk gene expression analysis Thomas

Processed data and normalized in Bioconductor by RMA **(Irizarry et al., 2003)**, which includes global background adjustment and quantile normalization. Samples from phase1 subjects in the antibiotics and control arm of the study were selected and statistical tests and correlation analyses were performed using MATLAB. Test details and significance cutoffs are reported in figure legends.

#### Luminex analysis

Statistical analysis was conducted in R (v 4.0.2) **(R Core Team, 2020)**. First, MFI data was log2 transformed and average MFI and CV was calculated from duplicate cultures where available. For samples with CV > 0.25, the duplicate that was closer to the average of all samples of that subject was kept and the other discarded. In case no other sample was available and CV > 0.25, the sample was discarded. Wells without indication of cytokine production were excluded. Statistical tests, correlation analysis, and hierarchical clustering were performed using the R packages stats (v 4.0.2), ggpubr (v 0.4.0) and pheatmap (v 1.0.12). Test details and statistical cutoffs are reported in the figure legends.

#### Bulk ATAC-seq pre-processing

Analysis of ATAC-seq data was very similar to the analysis of ChIP-Seq data. Reads were aligned using the BWA algorithm (mem mode; default settings; v 0.7.12) **(Li and Durbin, 2009)**. Duplicate reads were removed, only reads mapping as matched pairs and only uniquely mapped reads (mapping quality >= 1) were used for further analysis. Alignments were extended in silico at their 3’-ends to a length of 200 bp and assigned to 32-nt bins along the genome. The resulting histograms (genomic “signal maps”) were stored in bigWig files. Peaks were identified using the MACS algorithm (v 2.1.0) **(Zhang et al., 2008)** at a cutoff of p-value 1e-7, without control file, and with the –nomodel option. Peaks that were on the ENCODE blacklist of known false ChIP-Seq peaks were removed. Signal maps and peak locations were used as input data to Active Motif’s proprietary analysis program, which creates Excel tables containing detailed information on sample comparison, peak metrics, peak locations and gene annotations. For differential analysis, reads were counted in all merged peak regions (using Subread), and the replicates for each condition were compared using DESeq2 (v 1.24.0) **(Love et al., 2014)**.

#### Bulk ATAC-seq analysis

Quality control analysis of ATAC-seq data was performed using Rockefeller University workshop on analysis of ATAC-seq data in R and Bioconductor (https://rockefelleruniversity.github.io/RU_ATAC_Workshop.html). Of 57 unique samples processed, 51 passed QC criteria and, on average, we detected more than 42,000 genomic regions and more than 15×10^6^ unique ATAC tags per sample while the average fraction of reads in peaks was larger than 35% (**DataS3**). Passed samples showed the characteristic fragment length and TSS enrichment distribution (**DataS3**). DARs were annotated as promoter, distal and trans regulatory peak for a particular gene based on the distance from the middle of the peak to the nearest transcription start site (TSS) using the ChIPpeakAnno package in R (v. 3.24.1). Promoter, distal and trans regulatory peaks were defined as -2000 bp to +500 bp, -10kbp to +10kbp – promoter, and < -10kbp or > +10kbp from TSS, respectively. The hypergeometric distribution-based enrichment analysis was performed to identify the significance of the DARs. Reactome pathways and TF-target relationship using Chip-seq data from ENCODE (both downloaded from https://maayanlab.cloud/chea3/) were used to identify overrepresented pathways and TFs. EnrichmentMap Pipeline Collection (v 1.1.0) **(Merico et al., 2010)** for CytoScape (v 3.8.2) **(Shannon et al., 2003)** was used to create the pathway network. Significantly enriched Reactome pathways (p <= 0.05) for each genomic region were used as input. Pathways were clustered and annotated using the AutoAnnotate function within the pipeline. To test for enrichment of TF motifs in DARs, the chromVAR (v 1.8.0) **(Schep et al., 2017)** and motifmatchr (v 1.8.0) packages were used in R (v 3.6.0) **(R Core Team, 2020)**. In brief, TF motifs were downloaded from the JASPAR2016 core homo sapiens database **(Mathelier et al., 2016)** and merged regions were annotated for the presence of all TF binding motifs using the matchMotifs (motifmatchr) function with standard settings. Hypergeometric distribution-based enrichment analysis was then performed to identify enrichment of TF motifs in DARs. To determine the relationship between EpiTOF and ATAC-seq data, the Pearson correlation was computed between EpiTOF H3K27ac levels and normalized read counts in each merged peak region. Positively correlated merged peak regions with p-value < = 0.05 were selected for functional annotation. Enrichment analysis was performed as described above.

#### Bulk RNA-seq of purified immune cells

Alignment was performed using STAR version 2.7.3a **(Dobin et al., 2013)** and transcripts were annotated using GRCh38 Ensembl release 100. Transcript abundance estimates were calculated internal to the STAR aligner using the algorithm of htseq-count **(Anders et al., 2015)**. DESeq2 version 1.26.0 **(Love et al., 2014)** was used for differential expression analysis using the Wald test with a paired design formula and using its standard library size normalization.

#### Analysis of bulk transcriptomics data from previous TIV studies

Processed bulk transcriptomics data from nine independent TIV studies conducted between 2007 and 2012 were obtained from GEO (accessions: GSE47353, GSE59635, GSE29619, GSE74813, GSE59654, GSE59743, GSE74811, GSE29617, GSE74816) **(Barrett et al., 2013; Mohanty et al., 2015; Nakaya et al., 2011, 2015; Thakar et al., 2015; Tsang et al., 2014)**. After removing samples and genes with missing values as well as extraordinary vaccine time points, we selected only samples from subjects matching the same age range as the current study: 18 – 45 years of age. The remaining samples were batch corrected using ComBat from the sva package in R (v 3.36.0) with study as batch, no covariates, and otherwise standard settings. Statistical tests were performed using the R base and ComplexHeatmap (v 2.4.3) packages. Test details and statistical cutoffs are reported in the figure legends.

#### scATAC analysis

The CellRanger-atac pipeline (v1.1.0) by 10X Genomics was used for alignment (GRCh38 reference genome), de-duplication, and identification of cut sites for each sample. The samples were then combined using the CellRanger-atac aggregation procedure without depth normalization (--normalize=none). The resulting fragment file was read into SnapATAC **(Fang et al., 2020)**. SnapATAC was used to bin the genome (bin size of 5K) and create a cell-by-bin count matrix. Cells were identified as barcodes with at latest 1000 UMIs, and a promotor ratio (defined as: (fragments in promoter regions + 1) / (total fragments + 1)) of at least 0.1, resulting in a total of [state total number of cells in each experiment], as stated in the results section. Bins that mapped to chrY, mitochondrial DNA, or bins that overlap with ENCODE blacklist regions **(Amemiya et al., 2019)**, were removed. The remaining bins were used for dimensionality reduction using Truncated SVD with the irlba R package **(Baglama et al., 2019)**, and the first 50 dimensions were then used for clustering. MACS2 **(Zhang et al., 2008)** was then used to call peaks within each cluster using recommended parameters for ATACseq data (*--nomodel --shift 100 --ext 200 --qval 5e-2 -B --SPMR*). The cluster-specific peaks were merged to a single combined set. SnapATAC was then used to map the fragments to the combined peaks set and create a peak-by-cell binary matrix. In the H5N1/H5N1+AS03 dataset, deeply-sequenced libraries were downsampled to an average of 1500 fragments per barcode by randomly removing counts from these samples at a probability p=1500/(mean fragments per cell in the sample). The dimensionality reduction and clustering procedure described above was then repeated on the peak-by-cell matrix. ChromVAR **(Schep et al., 2017)** was used with default parameters and the JASPAR2016 **(Mathelier et al., 2016)** motif database to calculate motif accessibility scores and compute differentially accessible motifs in the data. Hotspot was used to identify informative gene modules that explain heterogeneity within the monocyte population **(DeTomaso and Yosef, 2020),** using the Bernoulli model and the top 2500 regions (ranked by highest autocorrelation z-score) for module calculation. Modules were then identified using the *create_modules* function, with *min_gene_threshold=200*. Similar modules were manually identified and merged by taking the average score across modules. Differentially accessible regions were identified using logistic regression with the *glm* function in R with the design: *y ∼ timepoint + donor + log_fragments* to control for donor and library size effects. The coefficient corresponding to the time point was then used as the logFC value, and a Wald test was used to get p-values. For numerical stability, we only included peaks that were detected in at least 5% of the cells included in each comparison. All custom scripts for preprocessing, correlation analysis, and differential accessibility analysis are posted in zenodo [doi: 10.5281/zenodo.4446316]. The hypergeometric distribution-based enrichment analysis was performed to identify the significance of the DARs (p <= 0.05 and detected in at least 5% of cells). Reactome pathways database (both downloaded from https://maayanlab.cloud/chea3/) were used to identify overrepresented pathways. Enrichr (Kuleshov et al., 2016) was used to conduct enrichment analysis of genomic regions within Hotspot modules 2, 3. Enrichr was also used to conduct enrichment analysis of DARs containing an IRF1 motif. Briefly, significant DARs (p <= 0.05 and detected in at least 5% of cells) carrying an IRF1 motif, as determined by chromVAR, were selected. Next, gene names with multiple associated DARs were collapsed in case all DARs changed in the same direction or otherwise discared. Subsequently, gene list was submitted to Enrichr for enrichment using the Reactome_2016 database. Similarly, we used Enrichr together with the ChEA_2016 databases to identify TF target genes enriched in genes that were enhanced after booster vaccination with H5N1+AS03 and that overlapped with changes in accessibility at promoter regions.

#### scRNA analysis

The CellRanger pipeline (v3.1.0) by 10X Genomics was used for alignment (GRCh38 reference genome), demultiplexing, cell-calling, and filtering. The filtered count matrices from each sample were then aggregated using the CellRanger aggregation procedure without depth normalization (--normalize=none). The resulting count matrix was analyzed with scVI (scvi-tools v0.7.1)**(Lopez et al., 2018)** with default hyperparameters to fit a low-dimensional latent space, using the experiment annotation for each sample as a batch label for batch correction. Visualization, clustering, and exploratory analyses were performed with VISION (v2.1.0)**(DeTomaso et al., 2019)**. Differential expression analysis between time points was performed with edgeR **(Robinson et al., 2010)** as described in the package documentation, using the *exactTest* hypothesis testing for each pairwise analysis.

#### Bulk transcriptomics vax010

Initial data quality was assessed by background level, 3’ labeling bias, and pairwise correlation among samples via the arrayQualityMetrics package in Bioconductor **(Kauffmann et al., 2009)**. CEL files were normalized via RMA **(Irizarry et al., 2003)**, which includes global background adjustment and quantile normalization. Probes mapping to multiple genes were discarded, and the remaining probes were collapsed to gene level by selecting the probe for each gene with the highest mean expression across all subjects. Statistical tests were performed in MATLAB and R.

## Supplementary Figure Legends

**Figure S1.**
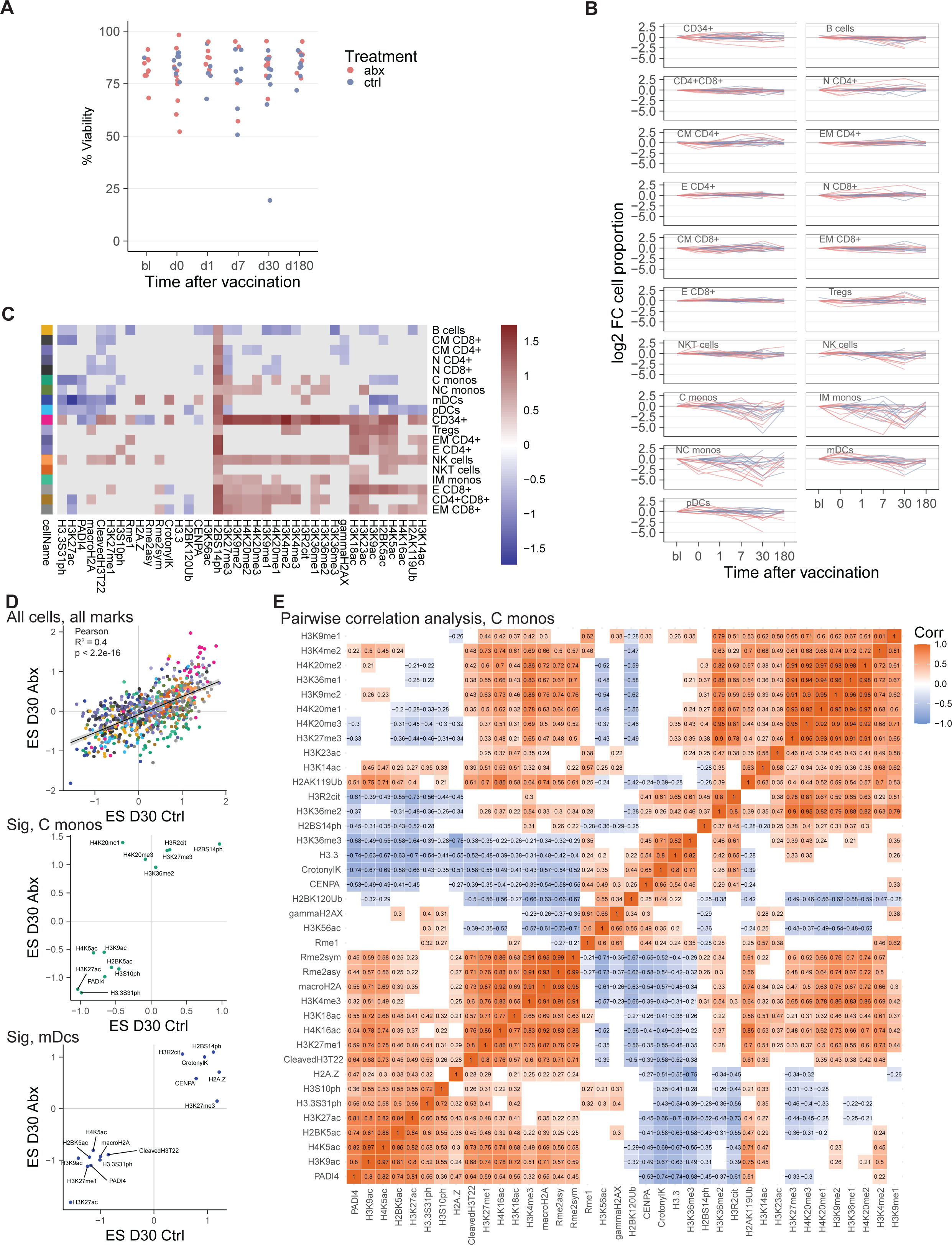
Cell type abundance and vaccine induced epigenomic changes by EpiTOF, related to Figure 1. (A) PBMC viability after thawing by vaccination time point. (B) Change in cell type abundance per subject. Wilcoxon signed rank test was used to compare changes at post-vaccine time points with d0 and p-values were corrected using the FDR approach. No comparison passed the threshold of fdr <= 0.05. (C) Heatmap showing histone modification changes at day 30 compared to day 0 in all detected immune cell subsets. Changes were calculated using the effect size approach. Only changes with an FDR <= 0.2 are shown. (D) Correlation of histone modification changes at day 30 compared to day 0 calculated separately for subjects in the control (x-axis) and antibiotics group (y-axis). For monocytes and mDCs, only significantly changed histone modifications are shown (FDR <= 0.2). (E) Correlation matrix showing the pair-wise correlation coefficient between all histone modification in classical monocytes.

**Figure S2.**
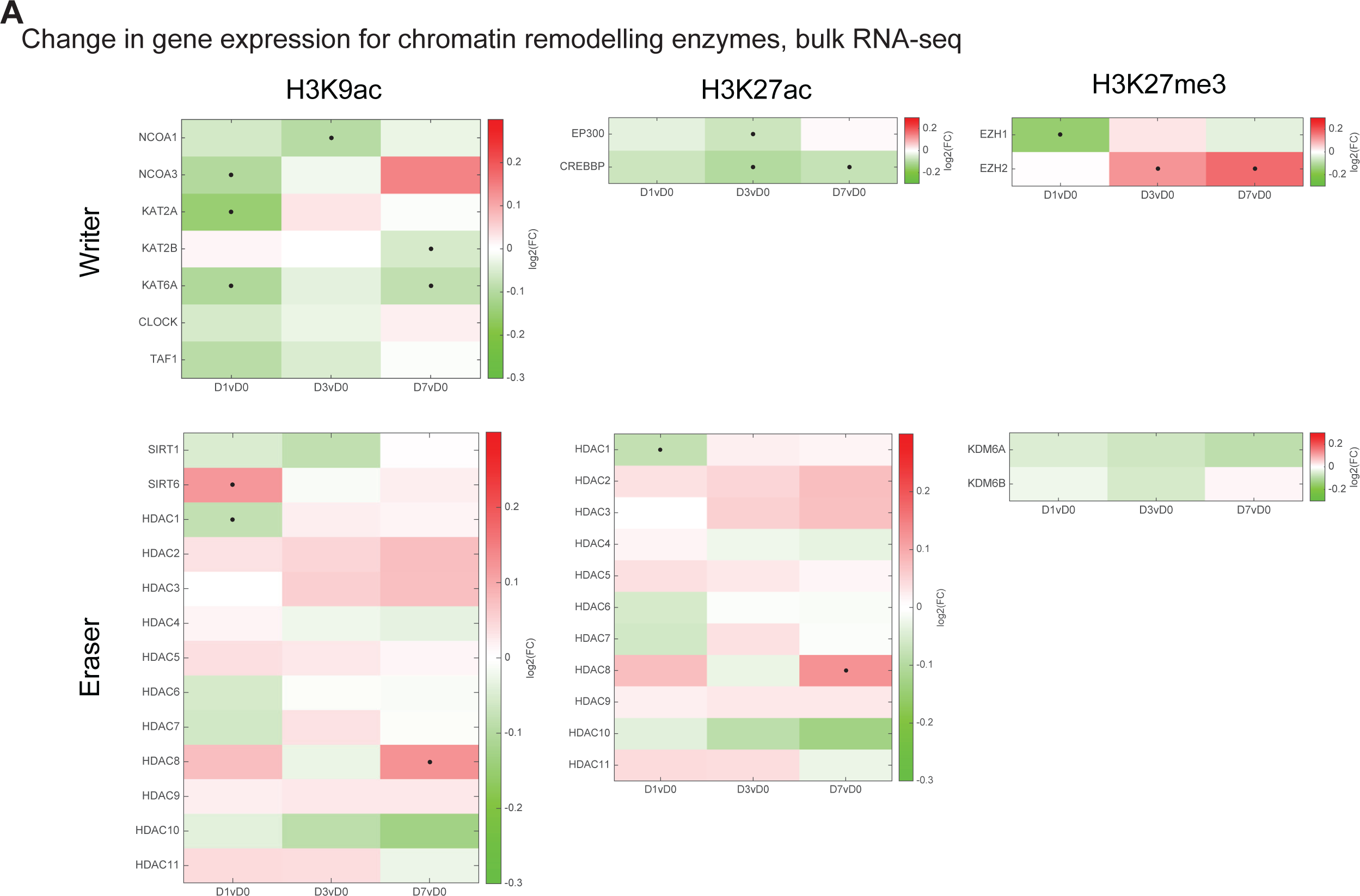
Analysis of vaccine-induced change in gene expression of histone modifying enzyme by bulk transcriptomics, related to Figure 1. (A) Heatmap showing the log2 fold change in gene expression relative to day 0 before vaccination. T-test was used for statistical testing. * p <= 0.05

**Figure S3.**
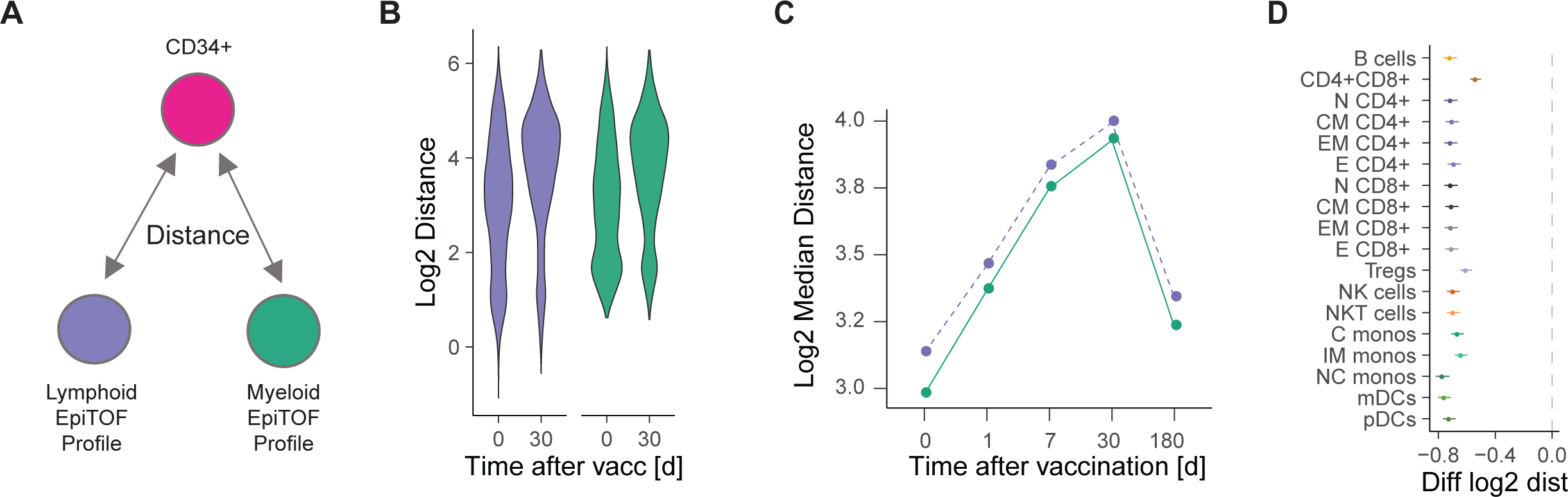
Histone modification profile distance of CD34^+^ progenitor cells by EpiTOF, related to Figure 1. (A) Cartoon of the analysis approach. The Euclidean distance between the histone modification profile of every single CD34^+^ progenitor cell to an average lymphoid or myeloid profile was calculated. (B) Violin plot showing the histone modification profile distance of single CD34^+^ progenitor cells to a common lymphoid (purple) or myeloid (turquoise) profile at the indicated time point using EpiTOF panel 2. (C) Median change in histone modification profile distance over time. (D) Change in histone modification profile distance of CD34^+^ progenitor cells to indicated cell types at day 30 after vaccination compared to day 0.

**Figure S4.**
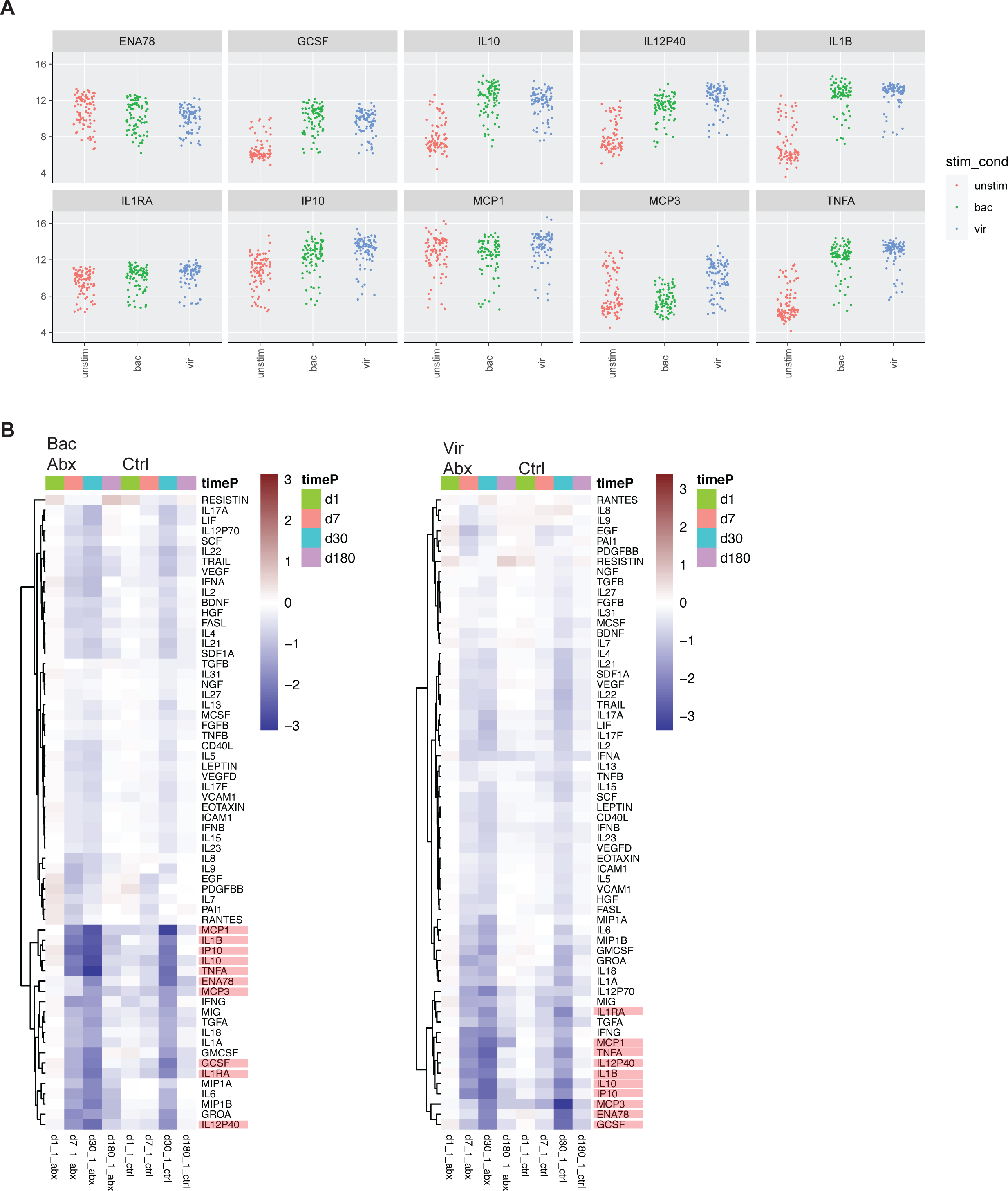
Cytokine production upon TLR stimulation, related to Figure 2. (A) Dot plot showing log2 cytokine levels in each TLR-stimulated PBMC culture by stimulation condition. (B) Heatmap showing the change in cytokine levels relative to day 0 separately for antibiotics-treated and control subjects.

**Figure S5.**
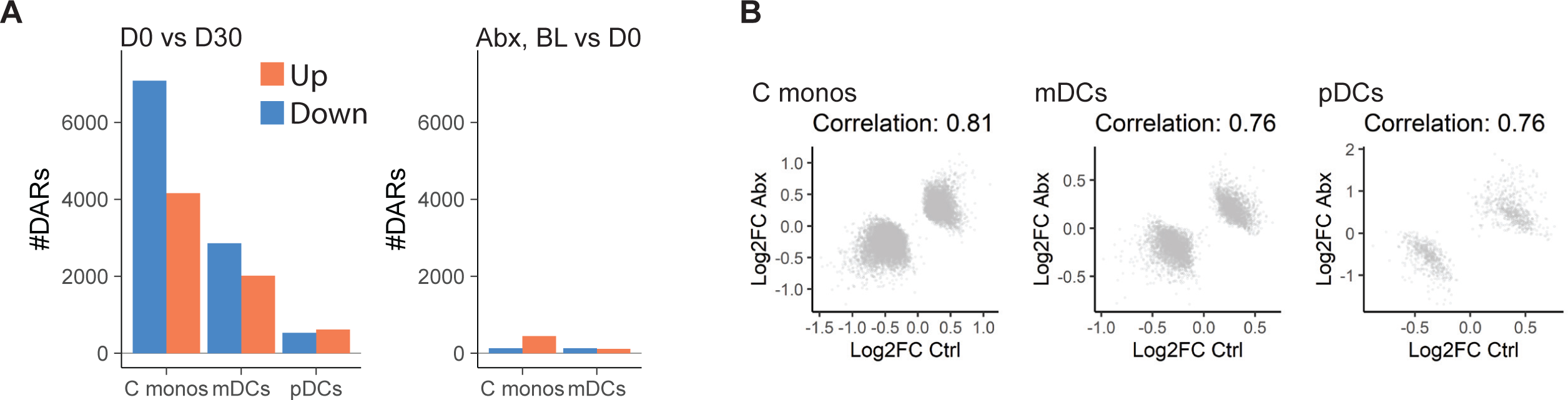
Vaccine-induced epigenomic changes by bulk ATAC-seq, related to Figure 3. (A) DARs at day 30 compared to day 0 (left) and day 0 vs baseline before antibiotics treatment (right, antibiotics subjects only). (B) DARs at day 30 compared to day 0 were calculated separately for control and antibiotics subjects. Log2 FC values from peaks that were significantly changed in the combined analysis (Figure 3b) were correlated with each using Pearson.

**Figure S6.**
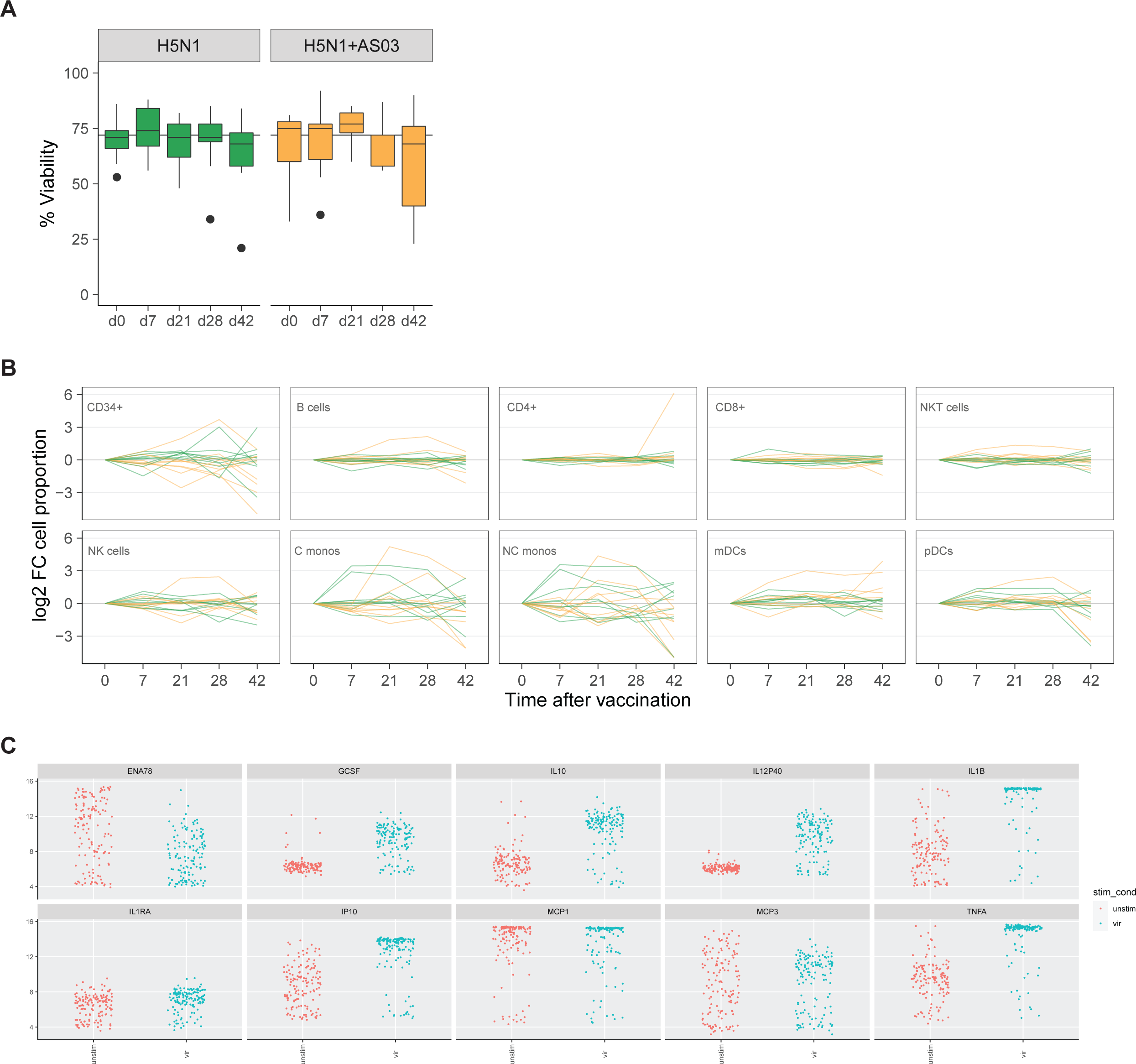
Changes in cell abundance and cytokine production upon TLR stimulation, related to Figure 5. (A) EpiTOF/Luminex PBMC viability after thawing by vaccination time point. (B) Change in cell type abundance per subject as measured by EPITOF. Wilcoxon signed rank test was used to compare changes at post-vaccine time points with d0 and p-values were corrected using the FDR approach. No comparison passed the threshold of fdr <= 0.05. (C) Dot plot showing log2 cytokine levels in each TLR-stimulated PBMC culture by stimulation condition.

**Figure S7.**
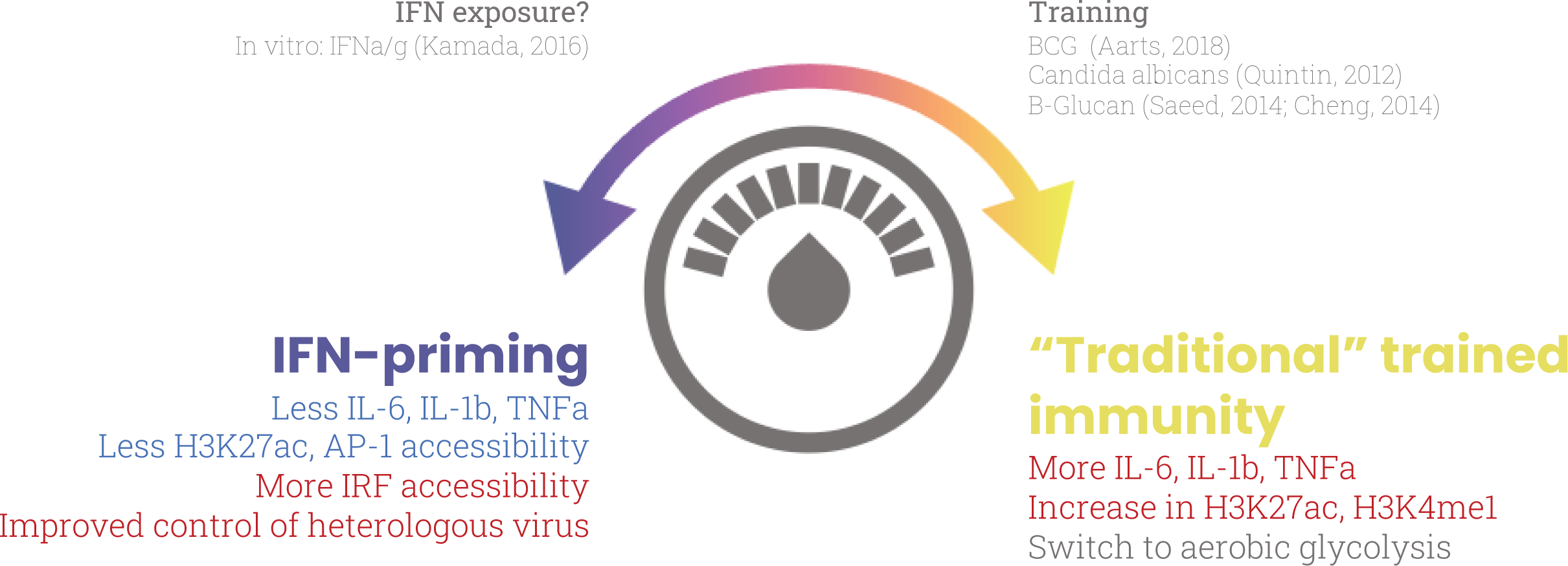
Model of bi-directional epigenomic reprogramming.

## References

Akondy, R.S., Fitch, M., Edupuganti, S., Yang, S., Kissick, H.T., Li, K.W., Youngblood, B.A., Abdelsamed, H.A., McGuire, D.J., Cohen, K.W., et al. (2017). Origin and differentiation of human memory CD8 T cells after vaccination. Nature 552, 362–367.

Alcántara-Hernández, M., Leylek, R., Wagar, L.E., Engleman, E.G., Keler, T., Marinkovich, M.P., Davis, M.M., Nolan, G.P., and Idoyaga, J. (2017). High-Dimensional Phenotypic Mapping of Human Dendritic Cells Reveals Interindividual Variation and Tissue Specialization. Immunity.

Allis, C.D., and Jenuwein, T. (2016). The molecular hallmarks of epigenetic control. Nat. Rev. Genet. 17, 487–500.

Amemiya, H.M., Kundaje, A., and Boyle, A.P. (2019). The ENCODE Blacklist: Identification of Problematic Regions of the Genome. Sci. Rep. 9, 9354.

Anders, S., Pyl, P.T., and Huber, W. (2015). HTSeq—a Python framework to work with high-throughput sequencing data. Bioinformatics 31, 166–169.

Arias, J., Alberts, A.S., Brindle, P., Claret, F.X., Smeal, T., Karin, M., Feramisco, J., and Montminy, M. (1994). Activation of cAMP and mitogen responsive genes relies on a common nuclear factor. Nature 370, 226–229.

Arts, R.J.W., Moorlag, S.J.C.F.M., Novakovic, B., Li, Y., Wang, S.-Y., Oosting, M., Kumar, V., Xavier, R.J., Wijmenga, C., Joosten, L.A.B., et al. (2018). BCG Vaccination Protects against Experimental Viral Infection in Humans through the Induction of Cytokines Associated with Trained Immunity. Cell Host Microbe 23, 89–100.e5.

Arunachalam, P.S., Wimmers, F., Mok, C.K.P., Perera, R.A.P.M., Scott, M., Hagan, T., Sigal, N., Feng, Y., Bristow, L., Tsang, O.T.-Y., et al. (2020). Systems biological assessment of immunity to mild versus severe COVID-19 infection in humans. Science.

Baglama, J., Reichel, L., and Lewis, B.W. (2019). irlba: Fast Truncated Singular Value Decomposition and Principal Components Analysis for Large Dense and Sparse Matrices.

Barrett, T., Wilhite, S.E., Ledoux, P., Evangelista, C., Kim, I.F., Tomashevsky, M., Marshall, K.A., Phillippy, K.H., Sherman, P.M., Holko, M., et al. (2013). NCBI GEO: archive for functional genomics data sets—update. Nucleic Acids Res. 41, D991–D995.

Barwick, B.G., Scharer, C.D., Bally, A.P.R., and Boss, J.M. (2016). Plasma cell differentiation is coupled to division-dependent DNA hypomethylation and gene regulation. Nat. Immunol. 17, 1216–1225.

Behre, G., Whitmarsh, A.J., Coghlan, M.P., Hoang, T., Carpenter, C.L., Zhang, D.-E., Davis, R.J., and Tenen, D.G. (1999). c-Jun Is a JNK-independent Coactivator of the PU.1 Transcription Factor. J. Biol. Chem. 274, 4939–4946.

Beisaw Arica, Kuenne Carsten, Guenther Stefan, Dallmann Julia, Wu Chi-Chung, Bentsen Mette, Looso Mario, and Stainier Didier Y.R. (2020). AP-1 Contributes to Chromatin Accessibility to Promote Sarcomere Disassembly and Cardiomyocyte Protrusion During Zebrafish Heart Regeneration. Circ. Res. 126, 1760–1778.

Benjamini, Y., and Hochberg, Y. (1995). Controlling the False Discovery Rate: A Practical and Powerful Approach to Multiple Testing. J. R. Stat. Soc. Ser. B Methodol. 57, 289–300.

Biddie, S.C., John, S., Sabo, P.J., Thurman, R.E., Johnson, T.A., Schiltz, R.L., Miranda, T.B., Sung, M.-H., Trump, S., Lightman, S.L., et al. (2011). Transcription Factor AP1 Potentiates Chromatin Accessibility and Glucocorticoid Receptor Binding. Mol. Cell 43, 145–155.

Bruggen, T. van der, Nijenhuis, S., Raaij, E. van, Verhoef, J., and Asbeck, B.S. van (1999). Lipopolysaccharide-Induced Tumor Necrosis Factor Alpha Production by Human Monocytes Involves the Raf-1/MEK1-MEK2/ERK1-ERK2 Pathway. Infect. Immun. 67, 3824–3829.

Buenrostro, J.D., Corces, M.R., Lareau, C.A., Wu, B., Schep, A.N., Aryee, M.J., Majeti, R., Chang, H.Y., and Greenleaf, W.J. (2018). Integrated Single-Cell Analysis Maps the Continuous Regulatory Landscape of Human Hematopoietic Differentiation. Cell 173, 1535–1548.e16.

Burny, W., Callegaro, A., Bechtold, V., Clement, F., Delhaye, S., Fissette, L., Janssens, M., Leroux-Roels, G., Marchant, A., van den Berg, R.A., et al. (2017). Different Adjuvants Induce Common Innate Pathways That Are Associated with Enhanced Adaptive Responses against a Model Antigen in Humans. Front. Immunol. 8.

Cao, R., and Zhang, Y. (2004). The functions of E(Z)/EZH2-mediated methylation of lysine 27 in histone H3. Curr. Opin. Genet. Dev. 14, 155–164.

Centers for Disease Control and Prevention (2020). CDC Seasonal Flu Vaccine Effectiveness Studies.

Cheung, P., Vallania, F., Warsinske, H.C., Donato, M., Schaffert, S., Chang, S.E., Dvorak, M., Dekker, C.L., Davis, M.M., Utz, P.J., et al. (2018). Single-Cell Chromatin Modification Profiling Reveals Increased Epigenetic Variations with Aging. Cell.

Cheung, W.L., Ajiro, K., Samejima, K., Kloc, M., Cheung, P., Mizzen, C.A., Beeser, A., Etkin, L.D., Chernoff, J., Earnshaw, W.C., et al. (2003). Apoptotic Phosphorylation of Histone H2B Is Mediated by Mammalian Sterile Twenty Kinase. Cell 113, 507–517.

Cho, K.-M., Kim, M.S., Jung, H.-J., Choi, E.-J., and Kim, T.S. (2019). Mst1-Deficiency Induces Hyperactivation of Monocyte-Derived Dendritic Cells via Akt1/c-myc Pathway. Front. Immunol. 10.

Cirovic, B., de Bree, L.C.J., Groh, L., Blok, B.A., Chan, J., van der Velden, W.J.F.M., Bremmers, M.E.J., van Crevel, R., Händler, K., Picelli, S., et al. (2020). BCG Vaccination in Humans Elicits Trained Immunity via the Hematopoietic Progenitor Compartment. Cell Host Microbe 28, 322–334.e5.

Corces, M.R., Buenrostro, J.D., Wu, B., Greenside, P.G., Chan, S.M., Koenig, J.L., Snyder, M.P., Pritchard, J.K., Kundaje, A., Greenleaf, W.J., et al. (2016). Lineage-specific and single-cell chromatin accessibility charts human hematopoiesis and leukemia evolution. Nat. Genet. 48, 1193–1203.

Corces, M.R., Trevino, A.E., Hamilton, E.G., Greenside, P.G., Sinnott-Armstrong, N.A., Vesuna, S., Satpathy, A.T., Rubin, A.J., Montine, K.S., Wu, B., et al. (2017). An improved ATAC-seq protocol reduces background and enables interrogation of frozen tissues. Nat. Methods 14, 959–962.

Das, M., Sabio, G., Jiang, F., Rincón, M., Flavell, R.A., and Davis, R.J. (2009). Induction of hepatitis by JNK-mediated expression of TNFα. Cell 136, 249–260.

DeTomaso, D., and Yosef, N. (2020). Identifying Informative Gene Modules Across Modalities of Single Cell Genomics. BioRxiv 2020.02.06.937805.

DeTomaso, D., Jones, M.G., Subramaniam, M., Ashuach, T., Ye, C.J., and Yosef, N. (2019). Functional interpretation of single cell similarity maps. Nat. Commun. 10, 4376.

Dobin, A., Davis, C.A., Schlesinger, F., Drenkow, J., Zaleski, C., Jha, S., Batut, P., Chaisson, M., and Gingeras, T.R. (2013). STAR: ultrafast universal RNA-seq aligner. Bioinformatics 29, 15–21.

Fang, R., Preissl, S., Li, Y., Hou, X., Lucero, J., Wang, X., Motamedi, A., Shiau, A.K., Zhou, X., Xie, F., et al. (2020). SnapATAC: A Comprehensive Analysis Package for Single Cell ATAC-seq. BioRxiv 615179.

Farlik, M., Halbritter, F., Müller, F., Choudry, F.A., Ebert, P., Klughammer, J., Farrow, S., Santoro, A., Ciaurro, V., Mathur, A., et al. (2016). DNA Methylation Dynamics of Human Hematopoietic Stem Cell Differentiation. Cell Stem Cell 19, 808–822.

Fontana, M.F., Baccarella, A., Pancholi, N., Pufall, M.A., Herbert, D.R., and Kim, C.C. (2015). JUNB Is a Key Transcriptional Modulator of Macrophage Activation. J. Immunol. 194, 177– 186.

Fujioka, S., Niu, J., Schmidt, C., Sclabas, G.M., Peng, B., Uwagawa, T., Li, Z., Evans, D.B., Abbruzzese, J.L., and Chiao, P.J. (2004). NF-1B and AP-1 Connection: Mechanism of NF-1B-Dependent Regulation of AP-1 Activity. MOL CELL BIOL 24, 14.

van Furth, R., and Cohn, Z.A. (1968). THE ORIGIN AND KINETICS OF MONONUCLEAR PHAGOCYTES. J. Exp. Med. 128, 415–435.

Garçon, N., Vaughn, D.W., and Didierlaurent, A.M. (2012). Development and evaluation of AS03, an Adjuvant System containing α-tocopherol and squalene in an oil-in-water emulsion. Expert Rev. Vaccines 11, 349–366.

Gaucher, D., Therrien, R., Kettaf, N., Angermann, B.R., Boucher, G., Filali-Mouhim, A., Moser, J.M., Mehta, R.S., Drake, D.R., Castro, E., et al. (2008). Yellow fever vaccine induces integrated multilineage and polyfunctional immune responses. J. Exp. Med. 205, 3119–3131.

Guilliams, M., Mildner, A., and Yona, S. (2018). Developmental and Functional Heterogeneity of Monocytes. Immunity 49, 595–613.

Hagan, T., Nakaya, H.I., Subramaniam, S., and Pulendran, B. (2015). Systems vaccinology: Enabling rational vaccine design with systems biological approaches. Vaccine 33, 5294–5301.

Hagan, T., Cortese, M., Rouphael, N., Boudreau, C., Linde, C., Maddur, M.S., Das, J., Wang, H., Guthmiller, J., Zheng, N.-Y., et al. (2019). Antibiotics-Driven Gut Microbiome Perturbation Alters Immunity to Vaccines in Humans. Cell 178, 1313–1328.e13.

Hannemann, N., Jordan, J., Paul, S., Reid, S., Baenkler, H.-W., Sonnewald, S., Bäuerle, T., Vera, J., Schett, G., and Bozec, A. (2017). The AP-1 Transcription Factor c-Jun Promotes Arthritis by Regulating Cyclooxygenase-2 and Arginase-1 Expression in Macrophages. J. Immunol. 198, 3605–3614.

Hedges, L.V., and Olkin, I. (2014). Statistical Methods for Meta-Analysis (Academic press).

Irizarry, R.A., Hobbs, B., Collin, F., Beazer-Barclay, Y.D., Antonellis, K.J., Scherf, U., and Speed, T.P. (2003). Exploration, normalization, and summaries of high density oligonucleotide array probe level data. Biostatistics 4, 249–264.

Kamath, A.T., Pooley, J., O’Keeffe, M.A., Vremec, D., Zhan, Y., Lew, A.M., D’Amico, A., Wu, L., Tough, D.F., and Shortman, K. (2000). The Development, Maturation, and Turnover Rate of Mouse Spleen Dendritic Cell Populations. J. Immunol. 165, 6762–6770.

Kamei, Y., Xu, L., Heinzel, T., Torchia, J., Kurokawa, R., Gloss, B., Lin, S.-C., Heyman, R.A., Rose, D.W., Glass, C.K., et al. (1996). A CBP Integrator Complex Mediates Transcriptional Activation and AP-1 Inhibition by Nuclear Receptors. Cell 85, 403–414.

Kauffmann, A., Gentleman, R., and Huber, W. (2009). arrayQualityMetrics—a bioconductor package for quality assessment of microarray data. Bioinformatics 25, 415–416.

Kaufmann, E., Sanz, J., Dunn, J.L., Khan, N., Mendonça, L.E., Pacis, A., Tzelepis, F., Pernet, E., Dumaine, A., Grenier, J.-C., et al. (2018). BCG Educates Hematopoietic Stem Cells to Generate Protective Innate Immunity against Tuberculosis. Cell 172, 176–190.e19.

Kazer, S.W., Aicher, T.P., Muema, D.M., Carroll, S.L., Ordovas-Montanes, J., Miao, V.N., Tu, A.A., Ziegler, C.G.K., Nyquist, S.K., Wong, E.B., et al. (2020). Integrated single-cell analysis of multicellular immune dynamics during hyperacute HIV-1 infection. Nat. Med.

Khurana, S., Coyle, E.M., Manischewitz, J., King, L.R., Gao, J., Germain, R.N., Schwartzberg, P.L., Tsang, J.S., and Golding, H. (2018). AS03-adjuvanted H5N1 vaccine promotes antibody diversity and affinity maturation, NAI titers, cross-clade H5N1 neutralization, but not H1N1 cross-subtype neutralization. Npj Vaccines 3, 1–12.

Kim, H.-J., and Bae, S.-C. Histone deacetylase inhibitors: molecular mechanisms of action and clinical trials as anti-cancer drugs. 14.

Kleinnijenhuis, J., Quintin, J., Preijers, F., Joosten, L.A.B., Ifrim, D.C., Saeed, S., Jacobs, C., van Loenhout, J., de Jong, D., Stunnenberg, H.G., et al. (2012). Bacille Calmette-Guerin induces NOD2-dependent nonspecific protection from reinfection via epigenetic reprogramming of monocytes. Proc. Natl. Acad. Sci. 109, 17537–17542.

Kotliarov, Y., Sparks, R., Martins, A.J., Mulè, M.P., Lu, Y., Goswami, M., Kardava, L., Banchereau, R., Pascual, V., Biancotto, A., et al. (2020). Broad immune activation underlies shared set point signatures for vaccine responsiveness in healthy individuals and disease activity in patients with lupus. Nat. Med. 26, 618–629.

Kuleshov, M.V., Jones, M.R., Rouillard, A.D., Fernandez, N.F., Duan, Q., Wang, Z., Koplev, S., Jenkins, S.L., Jagodnik, K.M., Lachmann, A., et al. (2016). Enrichr: a comprehensive gene set enrichment analysis web server 2016 update. Nucleic Acids Res. 44, W90–W97.

Kulis, M., Merkel, A., Heath, S., Queirós, A.C., Schuyler, R.P., Castellano, G., Beekman, R., Raineri, E., Esteve, A., Clot, G., et al. (2015). Whole-genome fingerprint of the DNA methylome during human B cell differentiation. Nat. Genet. 47, 746–756.

Langlais, D., Barreiro, L.B., and Gros, P. (2016). The macrophage IRF8/IRF1 regulome is required for protection against infections and is associated with chronic inflammationIRF8 and IRF1 regulate macrophage gene expression. J. Exp. Med. 213, 585–603.

Lasko, L.M., Jakob, C.G., Edalji, R.P., Qiu, W., Montgomery, D., Digiammarino, E.L., Hansen, T.M., Risi, R.M., Frey, R., Manaves, V., et al. (2017). Discovery of a selective catalytic p300/CBP inhibitor that targets lineage-specific tumours. Nature 550, 128–132.

Lee, Y.-H., Coonrod, S.A., Kraus, W.L., Jelinek, M.A., and Stallcup, M.R. (2005). Regulation of coactivator complex assembly and function by protein arginine methylation and demethylimination. Proc. Natl. Acad. Sci. U. S. A. 102, 3611–3616.

Li, H., and Durbin, R. (2009). Fast and accurate short read alignment with Burrows–Wheeler transform. Bioinformatics 25, 1754–1760.

Li, C., Bi, Y., Li, Y., Yang, H., Yu, Q., Wang, J., Wang, Y., Su, H., Jia, A., Hu, Y., et al. (2017a). Dendritic cell MST1 inhibits Th17 differentiation. Nat. Commun. 8, 14275.

Li, S., Sullivan, N.L., Rouphael, N., Yu, T., Banton, S., Maddur, M.S., McCausland, M., Chiu, C., Canniff, J., Dubey, S., et al. (2017b). Metabolic Phenotypes of Response to Vaccination in Humans. Cell 169, 862–877.e17.

Li, W., Xiao, J., Zhou, X., Xu, M., Hu, C., Xu, X., Lu, Y., Liu, C., Xue, S., Nie, L., et al. (2015). STK4 regulates TLR pathways and protects against chronic inflammation–related hepatocellular carcinoma. J. Clin. Invest. 125, 4239–4254.

Liu, Y., Lightfoot, Y.L., Seto, N., Carmona-Rivera, C., Moore, E., Goel, R., O’Neil, L., Mistry, P., Hoffmann, V., Mondal, S., et al. (2018). Peptidylarginine deiminases 2 and 4 modulate innate and adaptive immune responses in TLR-7–dependent lupus. JCI Insight 3.

Lopez, R., Regier, J., Cole, M.B., Jordan, M.I., and Yosef, N. (2018). Deep generative modeling for single-cell transcriptomics. Nat. Methods 15, 1053–1058.

Love, M.I., Huber, W., and Anders, S. (2014). Moderated estimation of fold change and dispersion for RNA-seq data with DESeq2. Genome Biol. 15, 1–21.

Mathelier, A., Fornes, O., Arenillas, D.J., Chen, C., Denay, G., Lee, J., Shi, W., Shyr, C., Tan, G., Worsley-Hunt, R., et al. (2016). JASPAR 2016: a major expansion and update of the open-access database of transcription factor binding profiles. Nucleic Acids Res. 44, D110–D115.

McElhaney, J.E., Beran, J., Devaster, J.-M., Esen, M., Launay, O., Leroux-Roels, G., Ruiz-Palacios, G.M., van Essen, G.A., Caplanusi, A., Claeys, C., et al. (2013). AS03-adjuvanted versus non-adjuvanted inactivated trivalent influenza vaccine against seasonal influenza in elderly people: a phase 3 randomised trial. Lancet Infect. Dis. 13, 485–496.

McInnes, L., Healy, J., and Melville, J. (2020). UMAP: Uniform Manifold Approximation and Projection for Dimension Reduction. ArXiv180203426 Cs Stat.

Meraz, M.A., White, J.M., Sheehan, K.C.F., Bach, E.A., Rodig, S.J., Dighe, A.S., Kaplan, D.H., Riley, J.K., Greenlund, A.C., Campbell, D., et al. (1996). Targeted Disruption of the Stat1 Gene in Mice Reveals Unexpected Physiologic Specificity in the JAK–STAT Signaling Pathway. Cell 84, 431–442.

Merico, D., Isserlin, R., Stueker, O., Emili, A., and Bader, G.D. (2010). Enrichment Map: A Network-Based Method for Gene-Set Enrichment Visualization and Interpretation. PLOS ONE 5, e13984.

Mitroulis, I., Ruppova, K., Wang, B., Chen, L.-S., Grzybek, M., Grinenko, T., Eugster, A., Troullinaki, M., Palladini, A., Kourtzelis, I., et al. (2018). Modulation of Myelopoiesis Progenitors Is an Integral Component of Trained Immunity. Cell 172, 147–161.e12.

Mohanty, S., Joshi, S.R., Ueda, I., Wilson, J., Blevins, T.P., Siconolfi, B., Meng, H., Devine, L., Raddassi, K., Tsang, S., et al. (2015). Prolonged Proinflammatory Cytokine Production in Monocytes Modulated by Interleukin 10 After Influenza Vaccination in Older Adults. J. Infect. Dis. 211, 1174–1184.

Monick, M.M., Carter, A.B., and Hunninghake, G.W. (1999). Human Alveolar Macrophages Are Markedly Deficient in REF-1 and AP-1 DNA Binding Activity. J. Biol. Chem. 274, 18075– 18080.

Nakashima, K., Hagiwara, T., Ishigami, A., Nagata, S., Asaga, H., Kuramoto, M., Senshu, T., and Yamada, M. (1999). Molecular Characterization of Peptidylarginine Deiminase in HL-60 Cells Induced by Retinoic Acid and 1α,25-Dihydroxyvitamin D3. J. Biol. Chem. 274, 27786–27792.

Nakaya, H.I., Wrammert, J., Lee, E.K., Racioppi, L., Marie-Kunze, S., Haining, W.N., Means, A.R., Kasturi, S.P., Khan, N., Li, G.-M., et al. (2011). Systems biology of vaccination for seasonal influenza in humans. Nat. Immunol. 12, 786–795.

Nakaya, H.I., Hagan, T., Duraisingham, S.S., Lee, E.K., Kwissa, M., Rouphael, N., Frasca, D., Gersten, M., Mehta, A.K., Gaujoux, R., et al. (2015). Systems Analysis of Immunity to Influenza Vaccination across Multiple Years and in Diverse Populations Reveals Shared Molecular Signatures. Immunity 43, 1186–1198.

Netea, M.G., Domínguez-Andrés, J., Barreiro, L.B., Chavakis, T., Divangahi, M., Fuchs, E., Joosten, L.A.B., van der Meer, J.W.M., Mhlanga, M.M., Mulder, W.J.M., et al. (2020). Defining trained immunity and its role in health and disease. Nat. Rev. Immunol. 20, 375–388.

Panda, D., Gjinaj, E., Bachu, M., Squire, E., Novatt, H., Ozato, K., and Rabin, R.L. (2019). IRF1 Maintains Optimal Constitutive Expression of Antiviral Genes and Regulates the Early Antiviral Response. Front. Immunol. 10.

Patel, A.A., Zhang, Y., Fullerton, J.N., Boelen, L., Rongvaux, A., Maini, A.A., Bigley, V., Flavell, R.A., Gilroy, D.W., Asquith, B., et al. (2017). The fate and lifespan of human monocyte subsets in steady state and systemic inflammation. J. Exp. Med. 214, 1913–1923.

Phanstiel, D.H., Van Bortle, K., Spacek, D., Hess, G.T., Shamim, M.S., Machol, I., Love, M.I., Aiden, E.L., Bassik, M.C., and Snyder, M.P. (2017). Static and Dynamic DNA Loops form AP-1-Bound Activation Hubs during Macrophage Development. Mol. Cell 67, 1037–1048.e6.

Pulendran, B., Li, S., and Nakaya, H.I. (2010). Systems Vaccinology. Immunity 33, 516–529.

Querec, T.D., Akondy, R.S., Lee, E.K., Cao, W., Nakaya, H.I., Teuwen, D., Pirani, A., Gernert, K., Deng, J., Marzolf, B., et al. (2009). Systems biology approach predicts immunogenicity of the yellow fever vaccine in humans. Nat. Immunol. 10, 116–125.

R Core Team (2020). R: A Language and Environment for Statistical Computing (Vienna, Austria: R Foundation for Statistical Computing).

Robinson, M.D., McCarthy, D.J., and Smyth, G.K. (2010). edgeR: a Bioconductor package for differential expression analysis of digital gene expression data. Bioinformatics 26, 139–140.

Saeed, S., Quintin, J., Kerstens, H.H.D., Rao, N.A., Aghajanirefah, A., Matarese, F., Cheng, S.- C., Ratter, J., Berentsen, K., van der Ent, M.A., et al. (2014). Epigenetic programming of monocyte-to-macrophage differentiation and trained innate immunity. Science 345, 1251086– 1251086.

Satpathy, A.T., Granja, J.M., Yost, K.E., Qi, Y., Meschi, F., McDermott, G.P., Olsen, B.N., Mumbach, M.R., Pierce, S.E., Corces, M.R., et al. (2019). Massively parallel single-cell chromatin landscapes of human immune cell development and intratumoral T cell exhaustion. Nat. Biotechnol. 37, 925–936.

Schep, A.N., Wu, B., Buenrostro, J.D., and Greenleaf, W.J. (2017). chromVAR: inferring transcription-factor-associated accessibility from single-cell epigenomic data. Nat. Methods 14, 975–978.

Schulte-Schrepping, J., Reusch, N., Paclik, D., Baßler, K., Schlickeiser, S., Zhang, B., Krämer, B., Krammer, T., Brumhard, S., Bonaguro, L., et al. (2020). Severe COVID-19 Is Marked by a Dysregulated Myeloid Cell Compartment. Cell S0092867420309922.

See, P., Dutertre, C.-A., Chen, J., Günther, P., McGovern, N., Irac, S.E., Gunawan, M., Beyer, M., Händler, K., and Duan, K. (2017). Mapping the human DC lineage through the integration of high-dimensional techniques. Science 356, eaag3009.

Shalek, A.K., Satija, R., Shuga, J., Trombetta, J.J., Gennert, D., Lu, D., Chen, P., Gertner, R.S., Gaublomme, J.T., Yosef, N., et al. (2014). Single-cell RNA-seq reveals dynamic paracrine control of cellular variation. Nature 510, 363–369.

Shannon, P., Markiel, A., Ozier, O., Baliga, N.S., Wang, J.T., Ramage, D., Amin, N., Schwikowski, B., and Ideker, T. (2003). Cytoscape: A Software Environment for Integrated Models of Biomolecular Interaction Networks. Genome Res. 13, 2498–2504.

Soares-Schanoski, A., Cruz, N.B., Castro-Jorge, L.A. de, Carvalho, R.V.H. de, Santos, C.A. dos, Rós, N. da, Oliveira, Ú., Costa, D.D., Santos, C.L.S. dos, Cunha, M. dos P., et al. (2019). Systems analysis of subjects acutely infected with the Chikungunya virus. PLOS Pathog. 15, e1007880.

Solier, S., and Pommier, Y. (2009). The apoptotic ring: a novel entity with phosphorylated histones H2AX and H2B and activated DNA damage response kinases. Cell Cycle Georget. Tex 8, 1853–1859.

Stender, J.D., Pascual, G., Liu, W., Kaikkonen, M.U., Do, K., Spann, N.J., Boutros, M., Perrimon, N., Rosenfeld, M.G., and Glass, C.K. (2012). Control of Proinflammatory Gene Programs by Regulated Trimethylation and Demethylation of Histone H4K20. Mol. Cell 48, 28– 38.

Sun, J.C., Lopez-Verges, S., Kim, C.C., DeRisi, J.L., and Lanier, L.L. (2011). NK Cells and Immune “Memory.” J. Immunol. 186, 1891–1897.

Tamura, T., Yanai, H., Savitsky, D., and Taniguchi, T. (2008). The IRF Family Transcription Factors in Immunity and Oncogenesis. Annu. Rev. Immunol. 26, 535–584.

Team, H.-C.S.P., and Consortium, H.-I. (2017). Multicohort analysis reveals baseline transcriptional predictors of influenza vaccination responses. Sci. Immunol. 2.

Thakar, J., Mohanty, S., West, A.P., Joshi, S.R., Ueda, I., Wilson, J., Meng, H., Blevins, T.P., Tsang, S., Trentalange, M., et al. (2015). Aging-dependent alterations in gene expression and a mitochondrial signature of responsiveness to human influenza vaccination. Aging 7, 38–52.

Tsai, E.Y., Falvo, J.V., Tsytsykova, A.V., Barczak, A.K., Reimold, A.M., Glimcher, L.H., Fenton, M.J., Gordon, D.C., Dunn, I.F., and Goldfeld, A.E. (2000). A Lipopolysaccharide-Specific Enhancer Complex Involving Ets, Elk-1, Sp1, and CREB Binding Protein and p300 Is Recruited to the Tumor Necrosis Factor Alpha Promoter In Vivo. Mol. Cell. Biol. 20, 6084– 6094.

Tsang, J.S., Schwartzberg, P.L., Kotliarov, Y., Biancotto, A., Xie, Z., Germain, R.N., Wang, E., Olnes, M.J., Narayanan, M., Golding, H., et al. (2014). Global Analyses of Human Immune Variation Reveal Baseline Predictors of Postvaccination Responses. Cell 157, 499–513.

Ventura, J.-J., Kennedy, N.J., Lamb, J.A., Flavell, R.A., and Davis, R.J. (2003). c-Jun NH2-Terminal Kinase Is Essential for the Regulation of AP-1 by Tumor Necrosis Factor. Mol. Cell. Biol. 23, 2871–2882.

Villani, A.-C., Satija, R., Reynolds, G., Sarkizova, S., Shekhar, K., Fletcher, J., Griesbeck, M., Butler, A., Zheng, S., Lazo, S., et al. (2017). Single-cell RNA-seq reveals new types of human blood dendritic cells, monocytes, and progenitors. Science 356, eaah4573.

Voss, A.K., Collin, C., Dixon, M.P., and Thomas, T. (2009). Moz and Retinoic Acid Coordinately Regulate H3K9 Acetylation, Hox Gene Expression, and Segment Identity. Dev. Cell 17, 674–686.

Vossenaar, E.R., Radstake, T.R.D.,Heijden, A. van der, Mansum, M.A.M. van, Dieteren, C., Rooij, D.-J. de, Barrera, P., Zendman, A.J.W., and Venrooij, W.J. van (2004). Expression and activity of citrullinating peptidylarginine deiminase enzymes in monocytes and macrophages. Ann. Rheum. Dis. 63, 373–381.

Weinert, B.T., Narita, T., Satpathy, S., Srinivasan, B., Hansen, B.K., Schölz, C., Hamilton, W.B., Zucconi, B.E., Wang, W.W., Liu, W.R., et al. (2018). Time-Resolved Analysis Reveals Rapid Dynamics and Broad Scope of the CBP/p300 Acetylome. Cell 174, 231–244.e12.

Wen, W., Zhu, F., Zhang, J., Keum, Y.-S., Zykova, T., Yao, K., Peng, C., Zheng, D., Cho, Y.-Y., Ma, W., et al. (2010). MST1 Promotes Apoptosis through Phosphorylation of Histone H2AX. J. Biol. Chem. 285, 39108–39116.

Wimmers, F., and Pulendran, B. (2020). Emerging technologies for systems vaccinology — multi-omics integration and single-cell (epi)genomic profiling. Curr. Opin. Immunol. 65, 57–64.

Wimmers, F., Subedi, N., van Buuringen, N., Heister, D., Vivié, J., Beeren-Reinieren, I., Woestenenk, R., Dolstra, H., Piruska, A., Jacobs, J.F.M., et al. (2018). Single-cell analysis reveals that stochasticity and paracrine signaling control interferon-alpha production by plasmacytoid dendritic cells. Nat. Commun. 9.

Yoshida, M., Kijima, M., Akita, M., and Beppu, T. (1990). Potent and specific inhibition of mammalian histone deacetylase both in vivo and in vitro by trichostatin A. J. Biol. Chem. 265, 17174–17179.

Youngblood, B., Hale, J.S., Kissick, H.T., Ahn, E., Xu, X., Wieland, A., Araki, K., West, E.E., Ghoneim, H.E., Fan, Y., et al. (2017). Effector CD8 T cells dedifferentiate into long-lived memory cells. Nature 552, 404–409.

Zanger, K., Radovick, S., and Wondisford, F.E. (2001). CREB Binding Protein Recruitment to the Transcription Complex Requires Growth Factor–Dependent Phosphorylation of Its GF Box. Mol. Cell 7, 551–558.

Zhang, Y., Liu, T., Meyer, C.A., Eeckhoute, J., Johnson, D.S., Bernstein, B.E., Nussbaum, C., Myers, R.M., Brown, M., Li, W., et al. (2008). Model-based Analysis of ChIP-Seq (MACS). Genome Biol. 9, R137.

Zhou, X., Li, W., Wang, S., Zhang, P., Wang, Q., Xiao, J., Zhang, C., Zheng, X., Xu, X., Xue, S., et al. (2019). YAP Aggravates Inflammatory Bowel Disease by Regulating M1/M2 Macrophage Polarization and Gut Microbial Homeostasis. Cell Rep. 27, 1176–1189.e5.

